# Lipoprotein(a) testing in Patients with Atherosclerotic Cardiovascular Disease in Five Large U.S. Health Systems

**DOI:** 10.1101/2024.03.07.24303948

**Authors:** Nishant P. Shah, Hillary Mulder, Betsy Lydon, Karen Chiswell, Xingdi Hu, Zachary Lampron, Lauren Cohen, Manesh R. Patel, Susan Taubes, Wenliang Song, Suresh R. Mulukutla, Anum Saeed, Daniel P. Morin, Steven M. Bradley, Adrian F. Hernandez, Neha J. Pagidipati

## Abstract

**Introduction:** Lipoprotein (a) [Lp(a)] is an independent, causal risk factor for atherosclerotic cardiovascular disease (ASCVD). However, testing patterns remain variable and it is unclear what patient level factors are associated with Lp(a) testing, and if testing changes clinical management.

**Methods:** A retrospective study using electronic medical record data from five health systems in the CardioHealth Alliance identified an ASCVD cohort that was divided into those with a Lp(a) test and those without a Lp(a) test between 2019-2021. Baseline characteristics and initiation of lipid lowering therapy (LLT) was assessed. Multivariable regression modeling was used to determine factors associated with Lp(a) testing.

**Results:** Among 595,684 ASCVD patients, only 2,587 (0.4%) were tested for Lp(a) between 2019 and 2021. In adjusted models, those who were older or Black were less likely to have Lp(a) testing, while those with familial hypercholesterolemia, ischemic stroke/TIA, PAD, prior LLT, or LDL-C ≥130mg/dL were more likely to be tested. Those with a Lp(a) test, regardless of the Lp(a) value, were more frequently initiated on any statin therapy (30.3% vs. 10.6%, p<0.001), ezetimibe (7.65 vs. 0.8%, p <0.001), or PCSK9i (6.7% vs. 0.3%, p <0.001) compared with those without a test. Those with an elevated Lp(a) level more frequently initiated ezetimibe (11.5% vs. 5.9%, P <0.001) or PCSK9i (10.9% vs. 4.8%, P <0.001), but not statin therapy.

**Conclusions:** Lp(a) testing in ASCVD patients is infrequent, with evidence of disparities among older or Black individuals. Testing for Lp(a), regardless of level, is associated with greater initiation of any LLT, while elevated Lp(a) is associated with greater initiation of non-statin LLT. Overall, initiation of LLT remained low, despite guideline indications for LLT in ASCVD patients. There is a critical need for multidisciplinary and inclusive approaches to raise awareness for Lp(a) testing and its implications on management.

## Introduction

Despite advancements in treatment, atherosclerotic cardiovascular disease (ASCVD) remains the leading cause of morbidity and mortality across the world^1,2^. Multiple societal guidelines recommend aggressive lowering of low-density lipoprotein cholesterol (LDL-C) in patients with ASCVD^3,4^, yet residual risk still remains. One major contributor to residual risk is elevated lipoprotein (a) [Lp(a)], an apolipoprotein B100 (apoB)-containing lipoprotein bound with apolipoprotein (a) [apo(a)] which has been shown to have an independent and causal effect on early onset atherosclerosis^5,6^. Lp(a) is primarily genetically determined^7,8^ and elevated levels affect one in five individuals^6^.

Current guideline recommendations regarding Lp(a) vary across societies. The American College of Cardiology/American Heart Association (ACC/AHA) recommend testing for Lp(a) in primary prevention individuals at borderline or intermediate risk in order to reclassify risk, and testing in all individuals (both primary and secondary prevention) with a premature family history of ASCVD^3^. The European Society of Cardiology/European Atherosclerosis Society (ESC/EAS) guidelines recommend measurement of Lp(a) once in a lifetime in all individuals to identify those with extremely elevated levels (>180mg/dL or >430 nmol/L) which could serve as an equivalent lifetime ASCVD risk to those with heterozygous familial hypercholesterolemia (FH)^4^. Additionally the ESC/EAS recommend testing in all individuals with a premature family history of ASCVD, and to reclassify risk in those at moderate and high risk^4^. Furthermore, the National Lipid Association (NLA)^9^ recommends testing for Lp(a) in those with first degree relatives with premature ASCVD, a personal history of premature ASCVD, or primary severe hypercholesterolemia.

Testing for patients with elevated Lp(a) is important for many reasons. First, elevated Lp(a) is considered a risk enhancer which may warrant aggressive LDL-C lowering therapy^3,4,9^. Second, given that Lp(a) production is primarily genetically mediated, elevated levels in patients could have implications for cascade screening of first degree relatives who may benefit from earlier preventive interventions^10,11^ Third, knowing an Lp(a) level can help determine how aggressive patients and clinicians need to be in optimizing modifiable cardiovascular risk factors in addition to LDL-C, such as diabetes, hypertension, and obesity.^10,12^. Additionally, there may be a role for aspirin therapy in primary prevention for patients with elevated Lp(a) if the bleeding risks are low^13,14^. Fourth, knowing an Lp(a) level may identify eligibility in currently enrolling clinical trials for therapies targeting Lp(a) ^15,16^ or may identify individuals who could benefit from future therapies, should they become available. Finally, in secondary prevention populations, higher Lp(a) levels predict subsequent cardiovascular events and may warrant aggressive combination lipid lowering therapy^17^.

However, it is unclear how often and what types of populations are being tested for Lp(a) in contemporary real-world clinical practice. The literature also remains limited on whether knowing an Lp(a) value has an impact on clinical management. To address these questions, we assessed data from five large health systems across the United States participating in the CardioHealth Alliance. The CardioHealth Alliance is a consortium established in 2021 with the goal of improving the implementation of evidence-based practices to improve the care and health of patients with cardiovascular, renal, and metabolic diseases (CardioHealthAlliance.org)

## Methods

A retrospective analysis was conducted using electronic health record (EHR) data from five large health systems within the CardioHealth Alliance, including Allina Health, Duke University Medical Center, University of Pittsburgh Medical Center, Vanderbilt University Medical Center, and Ochsner Health System. Each health system participated in the Patient-Centered Clinical Research Network (PCORnet©) and EHR data was mapped into a common data model for analysis. Data elements include diagnosis and procedure codes, laboratory data, demographics, health system encounters, and medication data.

For the current analysis we included adults ≥ 18 years of age with established ASCVD defined as acute coronary syndrome (ACS), (myocardial infarction and unstable angina), stable angina, transient ischemic attack (TIA), ischemic heart disease, peripheral arterial disease (PAD), or revascularization procedures (coronary revascularization such has percutaneous coronary interventions (PCI) or coronary artery bypass grafting (CABG), peripheral and cerebrovascular revascularization procedures). The diagnosis and procedure codes used for the analysis can be seen in the supplementary materials.

Patients must have also had an ASCVD event within 5 years prior to an index date. For patients with a Lp(a) test, the index date was defined as date of the first Lp(a) test result in 2019-2021. Patients without any Lp(a) test from 2015 to 2021 formed the non-Lp(a) test group. To permit comparisons of change in LLT following Lp(a) testing, the index date for this group was defined as the date of a randomly selected outpatient encounter in 2019-2021. Additionally, patients were required to have at least 2 encounters of any kind within the health system in the 2 years prior to the index date. The follow-up period ranged from the index date to 6 months after until the end of study period (6/30/2022).

Variables collected for the study included age, sex, race, ethnicity, vitals, medical history, medications, and laboratory data (Lp(a) level, LDL-C, total cholesterol, high density lipoprotein cholesterol (HDL-C), triglyceride levels, glycosylated hemoglobin (HbA1c), and creatinine). Lipid lowering therapy (LLT) was defined as either statin at any dose, high intensity statin (Rosuvastatin 20-40mg daily or Atorvastatin 40-80mg daily), monoclonal antibody based proprotein convertase substilisin/kexin type 9 inhibitors (PCSK9i), ezetimibe, bempedoic acid, bile acid sequestrants, and fenofibrates. Medical history, medication prescriptions, labs and vitals were assessed based on EHR records in the 12 months prior to the index date.

Baseline characteristics for the overall cohort, including demographics, medical history, lab values, and concomitant medications, were stratified by the presence and absence of an Lp(a) test result. Continuous variables were summarized as median (25^th^, 75^th^ percentile) and categorical variables were presented as frequencies (%). A robust Poisson multivariable regression model was used to determine factors associated with obtaining an Lp(a) test. Factors were pre-specified for inclusion in the model based on clinical relevance. Regression estimates were adjusted for site. Missing values for covariates that had ≤10% missingness were imputed via a single random draw from regression-based chained equations. Missingness >10% for important covariates resulted in the creation of a “Missing” category, and categorization of the continuous covariate when applicable. Relative risks with 95% confidence intervals (CI) and p-values were presented for all variables in the model except for site, in order to maintain site confidentiality. Continuous variables were checked for linearity with respect to the outcome; all relationships were linear.

Initiation of a medication was based on the presence of prescription within 6 months following the index date, and was assessed among patients without a corresponding prescription in the prior 12 months. Initiation of LLT within 6 months after the index date was described and compared for patients with vs without Lp(a) testing and for those with Lp(a) testing, elevated values (defined as Lp(a) ≥ 50mg/dL or ≥125 nmol/L) vs non-elevated, as well as by Lp(a) level (<50 mg/dL, 50-100 mg/dL, >100 mg/dL, among those with Lp(a) measured in mg/dL). P-values for comparisons between groups were calculated using Chi-squared or Fisher’s exact tests, as appropriate.

A two-sided p value of < 0.05 was considered nominally statistically significant. All analyses were performed by the Duke Clinical Research Institute (Durham, NC) using SAS v9.4 (SAS Institute, Inc. Cary, NC). The data were extracted through SAS queries that were distributed to health systems and executed against the most recent PCORnet <u>Common Data Model</u>. Limited data sets were delivered through site approved secure file transfer methods. The study was approved by the Duke University institutional review board under a waiver of HIPAA authorization and informed consent.

## Results

Among 707,212 ASCVD patients in the five health systems, 3,437 (0.5%) had an Lp(a) test between 2019 and 2021. Of the patients with an Lp(a) test, 2,781 had an ASCVD event or diagnosis in the 5 years prior to the Lp(a) test and at least 2 encounters in the health system 2 years prior to the Lp(a) test. After excluding patients with a prior Lp(a) test between 2015-2018, 2,587 (0.4% of the study cohort) remained in the Lp(a) testing cohort. For the cohort of patients without Lp(a) testing, 670,138 had at least 1 outpatient encounter between 2019 and 2021. Among these patients without Lp(a) testing, 593,097 had a diagnosis of ASCVD in 5 years prior to the outpatient encounter and at least 2 encounters in the health system in the 2 years prior to the outpatient encounter. This created an overall study cohort of 595,684 ASCVD patients active in the health systems with and without a Lp(a) test (Figure 1).

**Figure 1:**
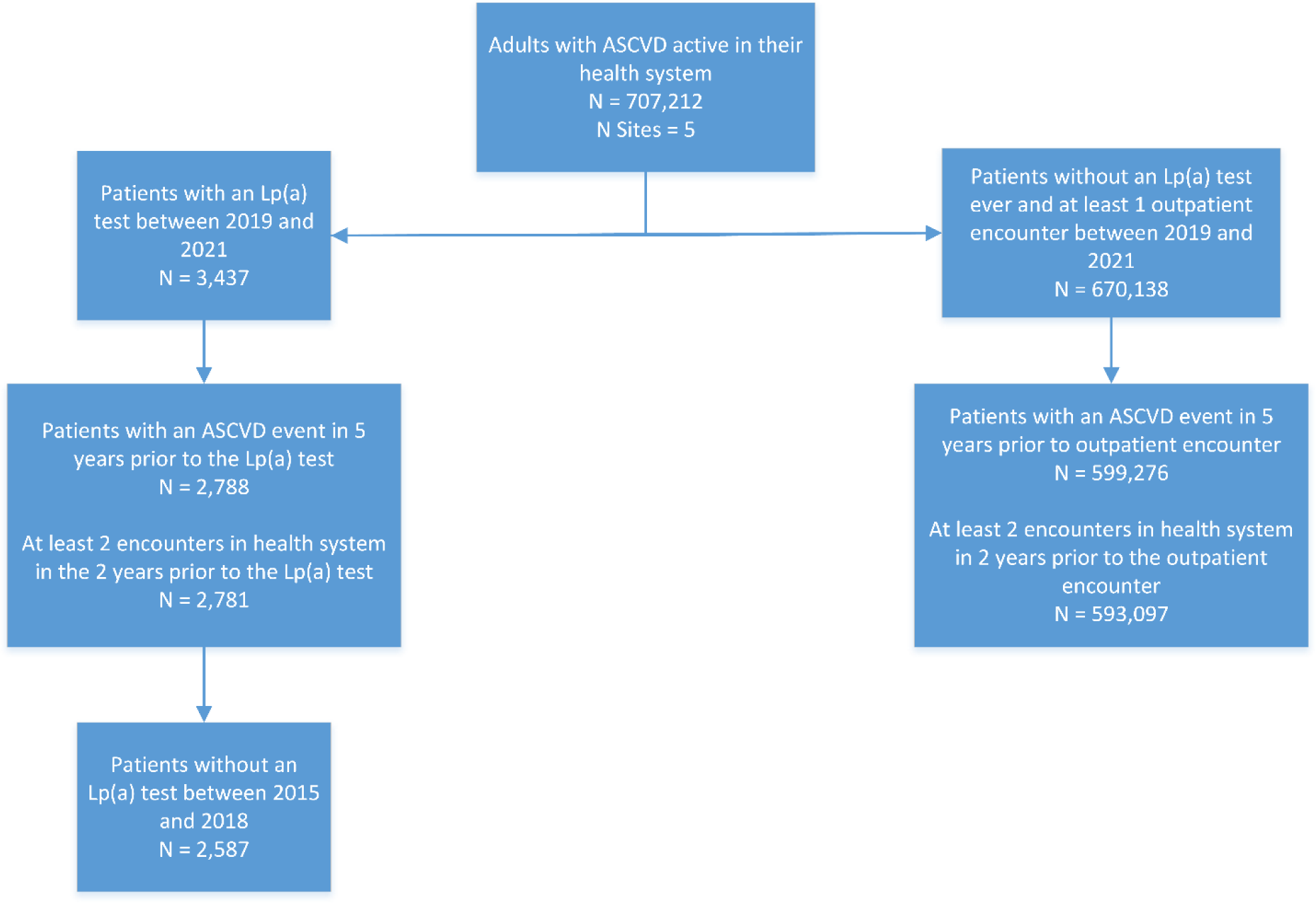
Consort diagram describing study cohort:

**Figure 2:**
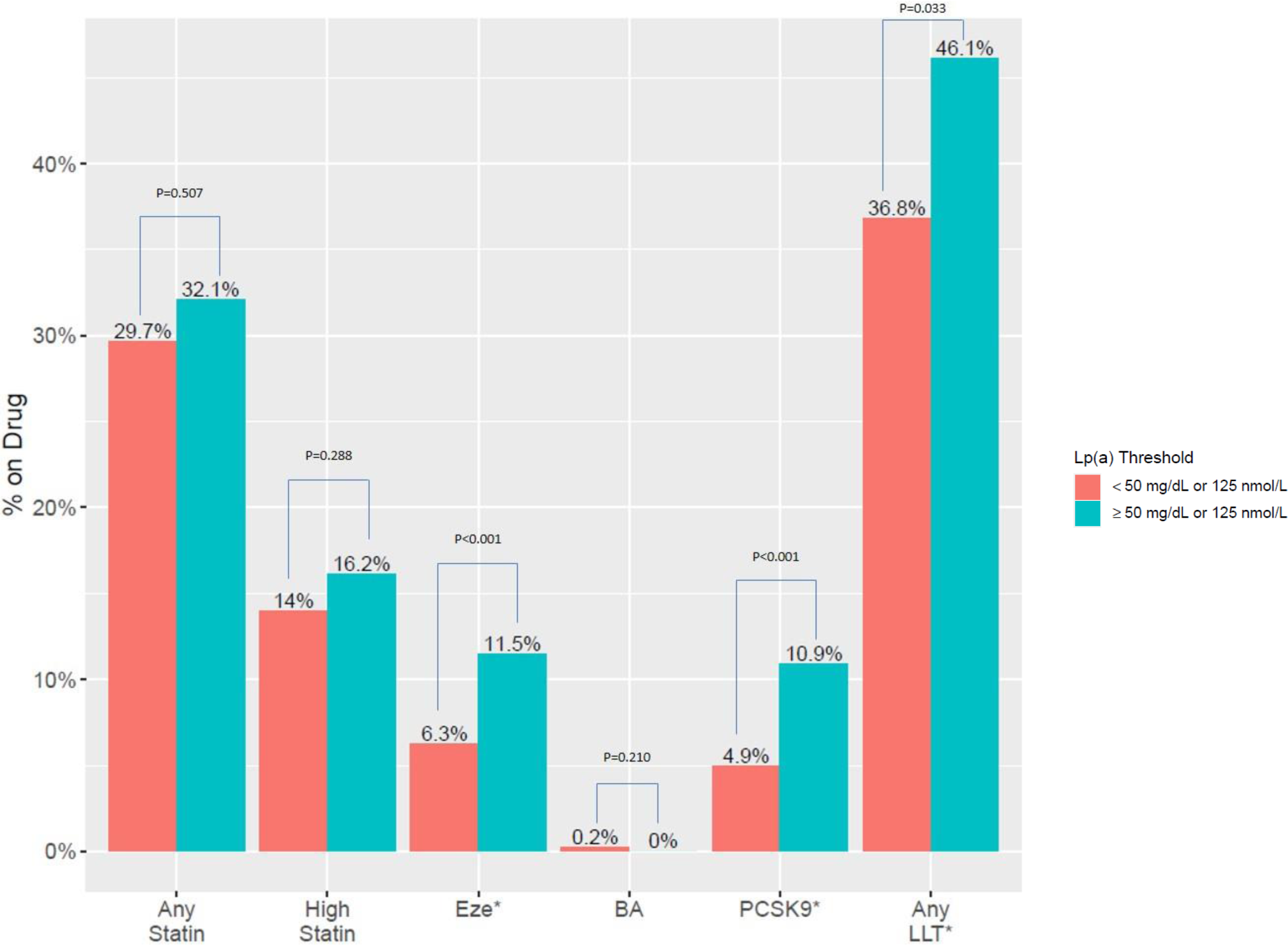
Initiation of Therapy During Follow Up among ASCVD patients with an Lp(a) test. Initiation defined by no prescription in the year prior to index date but a prescription is present in the 6 months following an Lp(a) test. Above Lp(a) threshold is defined as Lp(a) ≥ 50mg/dL or ≥125 nmol/L). Any LLT indicates a patient was initiated on one or more of the following prescriptions: statins, PCSK9i, ezetimibe, or bempedoic acid. BA: Bempedoic acid; Eze: ezetimibe; LLT: lipid lowering therapy; PCSK9: Monoclonal antibody based proprotein convertase subtilisin/kexin type 9 inhibitor;

Baseline characteristics of the overall study cohort stratified by presence or absence of Lp(a) testing are presented in Table 1. The median age of the overall cohort was 70 (Q1,Q3: 62,78) years. The cohort consisted of 45.0% female, and 84.5%, 13.0%, 1.4% White, Black, and Hispanic individuals, respectively. The majority of the study population had established coronary artery disease (75.4%), followed by peripheral arterial disease (31%), and stroke or TIA (20.4%). In terms of risk factors, 84.0% had hypertension, 78.0% had hyperlipidemia (with 3.7% diagnosed with FH), and 35.9% had diabetes mellitus. The median LDL-C was 81 (Q1,Q3:62,107) mg/dL, and only 48.9% of the study cohort were on any statin therapy. Furthermore, 3.0% were on ezetimibe, 0.9% were on PCSK9i, 1.8% were on fenofibrate, and <0.01% were on bempedoic acid.

**Table 1:**
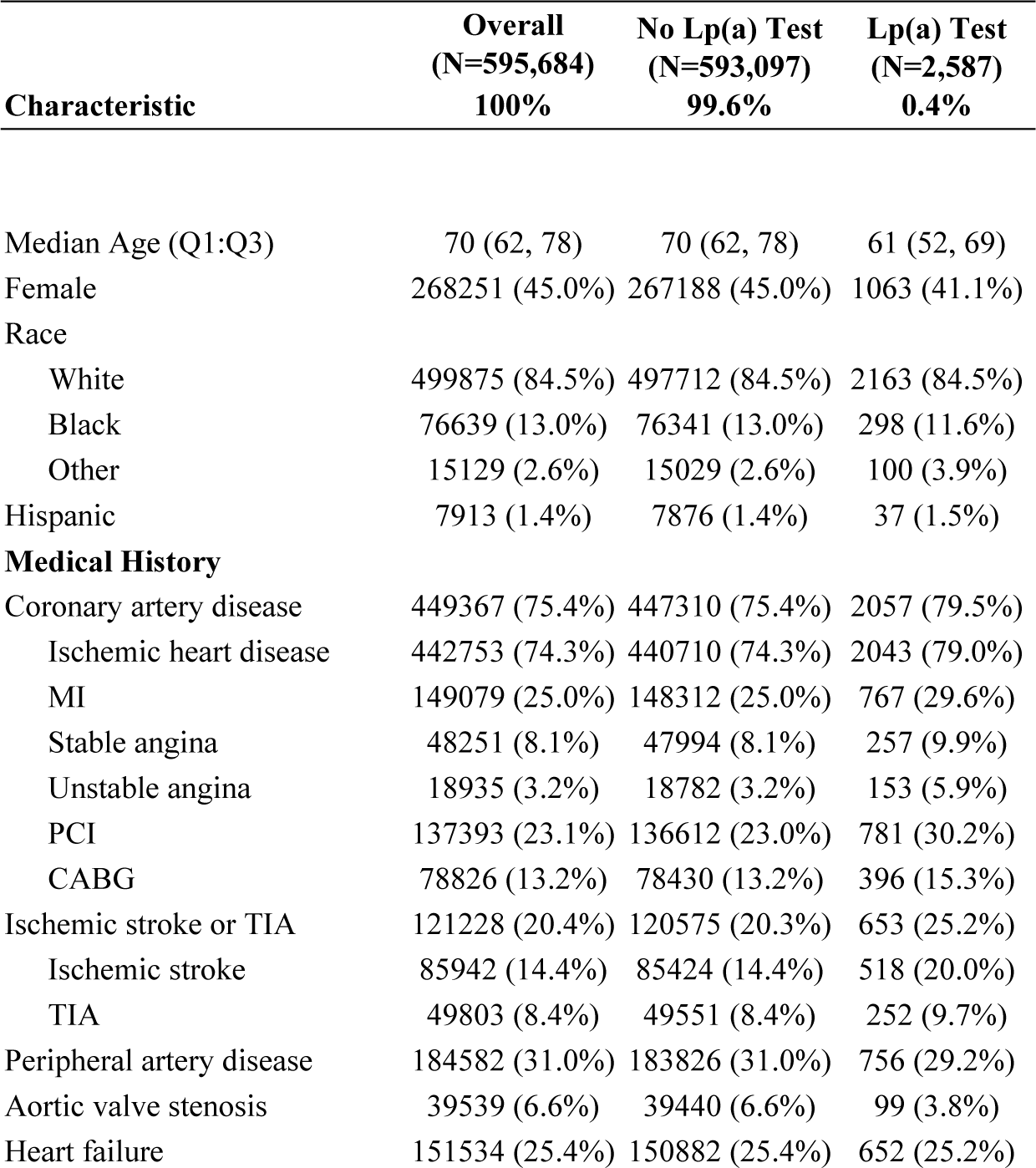
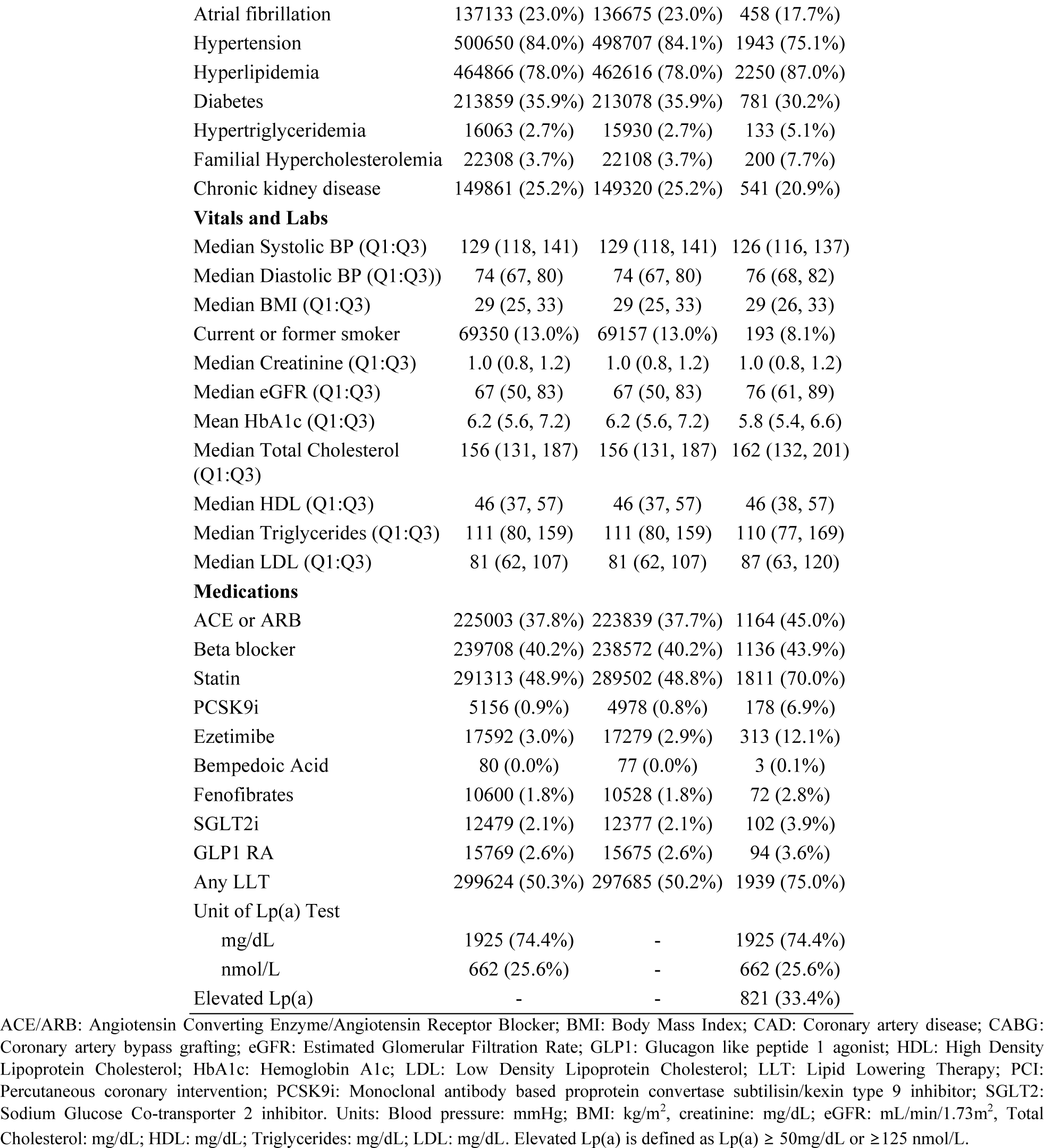
Baseline characteristics of ASCVD Patients with and without a Lp(a) Test.

The population with an Lp(a) test were younger (median age 61 vs 70 years), more frequently male (45.0% vs 41.1%), and less frequently Black (11.6% vs 13.0%) compared with patients without an Lp(a) test. In terms of medical co-morbidities, patients with an Lp(a) test had higher rates of ischemic stroke (25.2% vs 20.3%), hyperlipidemia (87.0% vs 78%), and FH (7.7% vs. 3.7%) compared with those without an Lp(a) test. In contrast, patients with an Lp(a) test had lower rates of atrial fibrillation (17.7% vs 23.0%), hypertension (75.1% vs 84.1%), and diabetes (30.2% vs 35.9%) compared with those without an Lp(a) test. LDL-C was higher in those with Lp(a) testing (87 vs. 81 mg/dL), and a greater proportion were on statin therapy (70% vs 48.8%). Additionally, more patients with an Lp(a) test were on ezetimibe (12.1% vs 2.9%) and PCSK9i (6.9% vs 0.8%) (Table 1).

Multivariable analysis suggested that several factors were independently associated with the likelihood of Lp(a) testing, including diagnosis of CAD, ischemic stroke/TIA, PAD, or heart failure (Figure 3, Supplemental Table 3). A diagnosis of FH or hyperlipidemia, along with use of any prior LLT, were also positively associated with Lp(a) testing. Of note, LDL-C level appeared to be associated with Lp(a) testing in a graded relationship, such that lower levels below 130 mg/dL were not associated with Lp(a) testing, but higher levels were increasingly associated with higher likelihood of receiving a Lp(a) test (Figure 3). In contrast, older age, Black race, higher BMI, current smoking status, and diagnosis of hypertension or diabetes were associated with a lower likelihood of Lp(a) testing. Missing HbA1c and lipid laboratory values in the 12 months prior to index date were also associated with lower likelihood of Lp(a) testing.

**Figure 3:**
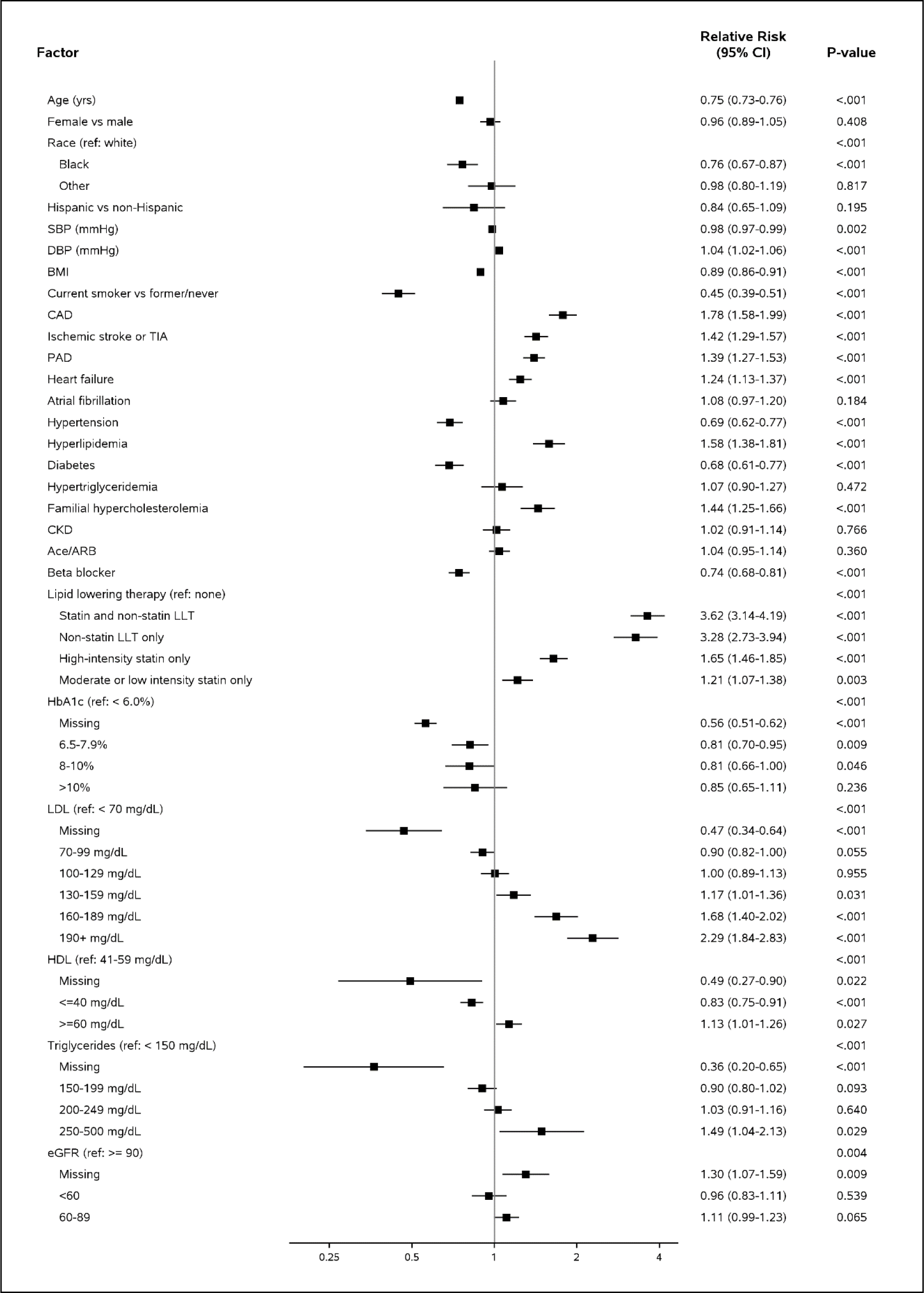
Forest plot of Multivariable Regression model describing factors associated with likelihood of testing for Lp(a.

The majority of Lp(a) testing was reported in mg/dL (74.4%) compared with nmol/L (25.6%). Those with elevated values, defined as Lp(a) ≥50mg/dL or ≥125 nmol/L, (n=821, 31.7%) were more frequently Black (18.2% vs. 8.6%), with higher LDL-C values (91 vs. 85 mg/dL) and higher rates of LLT use (78.1% vs. 73.5%) than those without elevated Lp(a) (Supplemental Table 1). Specifically, ezetimibe (16.3% vs. 10.1%) and PCSK9i (8.5% vs. 6.1%) were used at index more frequently in those with elevated Lp(a).

With regard to initiation of LLT, those with a Lp(a) test, regardless of the Lp(a) value, more often initiated LTT within 6 months of the index date (Table 2). Specifically, patients with a Lp(a) test were more frequently initiated on any statin therapy (30.3% vs. 10.6%, p<0.001), or high intensity statin therapy (14.6% vs. 5.1%, p<0.001) compared with those without a test. Initiation of PCSK9i (6.7% vs. 0.3%, p <0.001) and ezetimibe (7.65 vs. 0.8%, p <0.001) were more often in the group with Lp(a) testing as well.

**Table 2:** Initiation of lipid lowering therapy within 6 months after index date.

| LLT | Overall | No Lp(a) Test | Lp(a) Test | p-value |
| --- | --- | --- | --- | --- |
| Initiated Statins | 32493/304371 (10.7%) | 32258/303595 (10.6%) | 235/776 (30.3%) | <.001 |
| Initiated High Intensity Statins | 22631/441969 (5.1%) | 22422/440536 (5.1%) | 209/1433 (14.6%) | <.001 |
| Initiated PCSK9 | 1810/590528 (0.3%) | 1648/588119 (0.3%) | 162/2409 (6.7%) | <.001 |
| Initiated Ezetimibe | 4530/578092 (0.8%) | 4357/575818 (0.8%) | 173/2274 (7.6%) | <.001 |
| Initiated Bempedoic Acid | 57/595604 (0.0%) | 53/593020 (0.0%) | 4/2584 (0.2%) | -- |
| Initiated Any LLT | 33279/296060 (11.2%) | 33028/295412 (11.2%) | 251/648 (38.7%) | <.001 |

Initiation defined by no prescription in the year prior to index date but a prescription is present in the 6 months following index date. For patients with Lp(a) test, the index date was the date of the earliest Lp(a) test in 2021. For patients without an Lp(a) test the index date was the date of a randomly selected outpatient visit in 2021. Any LLT indicates a patient was initiated on one or more of the following prescriptions: statins, PCSK9i, ezetimibe, or bempedoic acid. LLT: lipid lowering therapy; PCSK9i: Monoclonal antibody based proprotein convertase subtilisin/kexin type 9 inhibitor

The Lp(a) value itself was associated with initiation of non-statin LLT (Figure 2). Among patients with Lp(a) values, there was no difference in the initiation of any statin (32.1% vs. 29.7%, p=0.51), high intensity statin (16.2% vs. 14.0%, p=0.29), or in up-titration from regular- to high-intensity statin dosing (49.3% vs. 41.1%, p-0.27) in those with elevated vs non-elevated Lp(a) levels (Table 3). However, initiation rates of PCSK9i (10.9% vs. 4.8%, P <0.001) and ezetimibe (11.5% vs. 5.9%, P <0.001) were significantly higher in those with elevated Lp(a) levels. Of note, there appeared to be a graded relationship such that those with highly elevated Lp(a) levels (≥ 100 mg/dL) were more likely to initiate PCSK9i or ezetimibe than those with Lp(a) 50-100 mg/dL or Lp(a) <50 mg/dL, respectively (PCSK9i: 15.5% vs 7.4% vs 5.5%, p<0.001; ezetimibe: 15.6% vs 12.8% vs 7.3%, p<0.001) (Supplemental Table 2). This trend was not present for statin initiation.

**Table 3:** Initiation of Lipid Lowering Therapy within 6 months after index date by Elevated Lp(a)

| LLT | Elevated Lp(a) |  |  | p-value |
| --- | --- | --- | --- | --- |
|  | Overall | No | Yes |  |
| Initiated Statins | 221/726 (30.4%) | 145/489 (29.7%) | 76/237 (32.1%) | 0.507 |
| Initiated High Intensity Statins | 198/1351 (14.7%) | 129/924 (14.0%) | 69/427 (16.2%) | 0.288 |
| Initiated PCSK9 | 158/2289 (6.9%) | 76/1538 (4.9%) | 82/751 (10.9%) | <.001 |
| Initiated Ezetimibe | 171/2153 (7.9%) | 92/1466 (6.3%) | 79/687 (11.5%) | <.001 |
| Initiated Bempedoic Acid | 4/2454 (0.2%) | 4/1634 (0.2%) | 0/820 (0.0%) | 0.210 |
| Initiated Any LLT | 238/601 (39.6%) | 155/421 (36.8%) | 83/180 (46.1%) | 0.033 |

Initiation defined by no prescription in the year prior to index date but a prescription is present in the 6 months following an Lp(a) test. Elevated Lp(a) threshold is defined as Lp(a) ≥ 50mg/dL or ≥ 125 nmol/L). Any LLT indicates a patient was initiated on one or more of the following prescriptions: statins, PCSK9i, ezetimibe, or bempedoic acid. LLT: lipid lowering therapy; PCSK9i: Monoclonal antibody based proprotein convertase subtilisin/kexin type 9 inhibitor

## Discussion

Across five large U.S. health systems within the CardioHealth Alliance, the frequency of Lp(a) testing was very low (0.4%) among patients with ASCVD. Disparities in Lp(a) testing were apparent, as older age, Black race, higher BMI, current smoking status, and diagnosis of hypertension or diabetes were associated with a lower likelihood of Lp(a) testing. Testing for Lp(a), regardless of Lp(a) level, was associated with greater initiation of LLT, including statin, ezetimibe, and PCSK9i. Interestingly, an elevated Lp(a) result was associated with ezetimibe and PCSK9i initiation, but not statin initiation or up-titration.

The low observed rate of Lp(a) testing among a secondary prevention population is consistent with previous reports. In a large claims data analysis performed on 4 million patient records in Germany, the frequency of Lp(a) testing was similarly low at 0.34%^18^. An analysis of U.S. claims data revealed that only 0.7% of secondary prevention patients were tested for Lp(a).^19^ Reasons behind the consistently low testing rates for Lp(a) in patients with ASCVD are likely multifactorial, and may include lack of clinician understanding or awareness of Lp(a) as a prevalent and causal risk factor for ASCVD^20^. Further, current clinical guidelines are inconsistent about recommendations for Lp(a) testing, though most recommend testing in high-risk patients^3,9^, if not all patients at least once in a lifetime^4^. In addition, there are no available therapies that are currently indicated to lower Lp(a), though several are in clinical trials (NCT04023552; NCT05581303)^21,22^. However, even in the absence of Lp(a)-lowering therapies, testing for Lp(a) is still clinically relevant for risk stratification, aggressive preventive management such as early use of statins, and testing of first-degree relatives.^6,11^

Those who did undergo Lp(a) testing in our study population were different in important ways from those who did not undergo testing, revealing potential disparities in testing patterns. Older age was associated with a lower likelihood of Lp(a) testing; this may be related to the NLA guidelines which specifically recommend Lp(a) testing for individuals with premature ASCVD, and who are therefore young by definition^9^. Black race was also associated with a 24% lower likelihood of Lp(a) testing; this likely represents a disparity in care, as Black individuals are known to have generally higher Lp(a) levels than White individuals, with correspondingly higher Lp(a)-related ASCVD risk^19,23^. As far as we are aware, this disparity in Lp(a) testing has not been shown before, and is important for addressing health equity in ASCVD prevention and management. Unsurprisingly, individuals with missing HbA1c and lipid levels were less likely to undergo Lp(a) testing, potentially because they were not receiving routine preventive care.

In contrast, several factors were associated with greater likelihood of Lp(a) testing, including prior CAD, ischemic stroke/TIA, PAD, or heart failure. This pattern is consistent with prior studies^18,24^ likely because of guideline recommendations for testing in high-risk populations^3,9^. Similarly, those with hyperlipidemia or FH were more likely to undergo testing, which also has been seen in prior studies and is consistent with guideline recommendations and the observation that elevated Lp(a) is more common in individuals with elevated LDL-C and/or FH^25,26^ Interestingly, LDL-C level appeared to have a graded relationship with Lp(a) testing, with increasing levels beyond 130 mg/dL being associated with greater likelihood of Lp(a) testing.

We also found that Lp(a) testing regardless of Lp(a) level, along with the level itself, are both associated with changes in LLT in the 6 months after testing. Lp(a) testing (compared with no testing) was associated with initiation of both statins and non-statin therapies, while an elevated Lp(a) level (compared with a non-elevated level) was associated only with initiation of PCSK9i and ezetimibe. These results suggest that clinicians may be acting on Lp(a) results with more aggressive therapies, indicating that testing may have an impact on clinical management (though causality cannot be established in this observational analysis). This is consistent with a prior study from Germany which showed increased treatment intensity after Lp(a) testing^18^; our study extends these results to the U.S. Further, our finding that statins were initiated regardless of Lp(a) result may reflect overall clinician concern and/or better care in those with an Lp(a) result, given that all of these individuals have a clear indication for statin therapy. Notably, however, only 30% of individuals with an Lp(a) test were initiated on a statin after testing, compared with 11% in those without testing, reflective of a continued gap in guideline-based care. In contrast with statins, PCSK9i and ezetimibe were more likely to be initiated in those with Lp(a) testing than in those without, but among those with testing, were only more likely to be initiated in those with an elevated Lp(a) level. This may indicate that clinicians are responding to the level itself by initiating more advanced LLT, which is consistent with guidelines that recommend more aggressive LDL-C lowering in those with elevated Lp(a)^3,9^. Again, however, the overall rate of initiation of these therapies was relatively low, with 11% and 12% of individuals with elevated Lp(a) initiating PCSK9i and ezetimibe, respectively.

Our study has several limitations that are important to note. First, this was a retrospective analysis that evaluated EHR data, so there is the potential for missing data, especially data generated outside the studied health systems; we attempted to minimize this risk by ensuring that all individuals in the study received regular care within these systems. Second, our study period overlapped with the advent of the COVID-19 pandemic period, which could have decreased the frequency of encounters and lab values. Third, adherence patterns and intolerance to LLT could not be captured, which could have influenced the distribution of various LLT medications.

## Conclusion

Across 5 large US health systems, the frequency of Lp(a) testing among patients with ASCVD remains low at 0.4%. Additionally, disparities exist as individuals who are older or Black were particularly unlikely to have a Lp(a) test. Those with Lp(a) testing were more likely to initiate any LLT regardless of Lp(a) level, including statins, while those with an elevated Lp(a) level were more likely to initiate PCSK9i or ezetimibe. However, the overall initiation of LLT remained low in this population, despite clear guidelines indications for LLT in patients with ASCVD. Thus, there is a critical need for multidisciplinary and inclusive approaches to raise awareness of Lp(a) testing and its implications for aggressive preventive management. Such awareness may increase if Lp(a)-lowering therapies, which are currently being tested, are shown to provide clinical benefit.

Disclosures:

The authors would like to provide the following disclosures: Disclosures: NPS: Research Grants: Amgen, Janssen, NIH; Consultant/Advisor: Esperion, Amgen, Norvartis; XH: Employed by Novartis Pharmaceuticals Corporation; MRP: Research grants: Amgen, Bayer, Janssen, Heartflow, NIH, Gordon and Betty Moore Foundation; Consultant/Advisor: Bayer, Janssen, Novartis, Amgen; ST: Employed by Novartis Pharmaceuticals Corporation; NJP: Research Grants: Amgen, AstraZeneca, Baseline Study LLC, Boehringer Ingleheim; Duke Clinical Research Institute, Eggland’s Best, Eli Lilly & Company, Novartis, Novo Nordisk Pharmaceutical Company, Sanofi-S.A, Verily Sciences Research Company; Consultant/Advisor: AstraZeneca; Boehringer Ingleheim; Esperion Therapeutics; Eli Lilly & Company; Novo Nordisk Pharmaceutical Company. All other authors do not have disclosures to report.

## Data Availability

Data will be made available upon request

---

**Supplementary Table 1:**
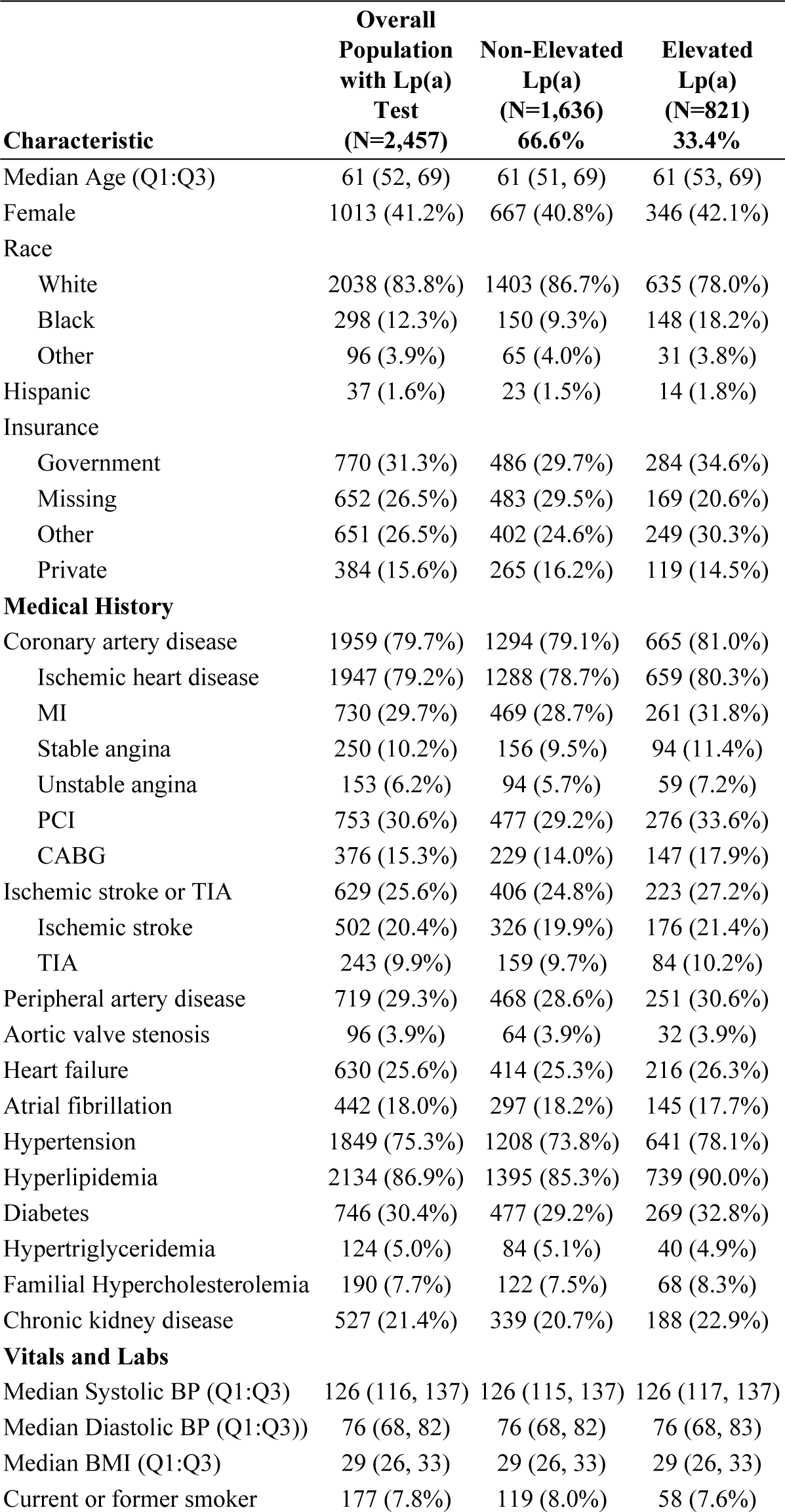
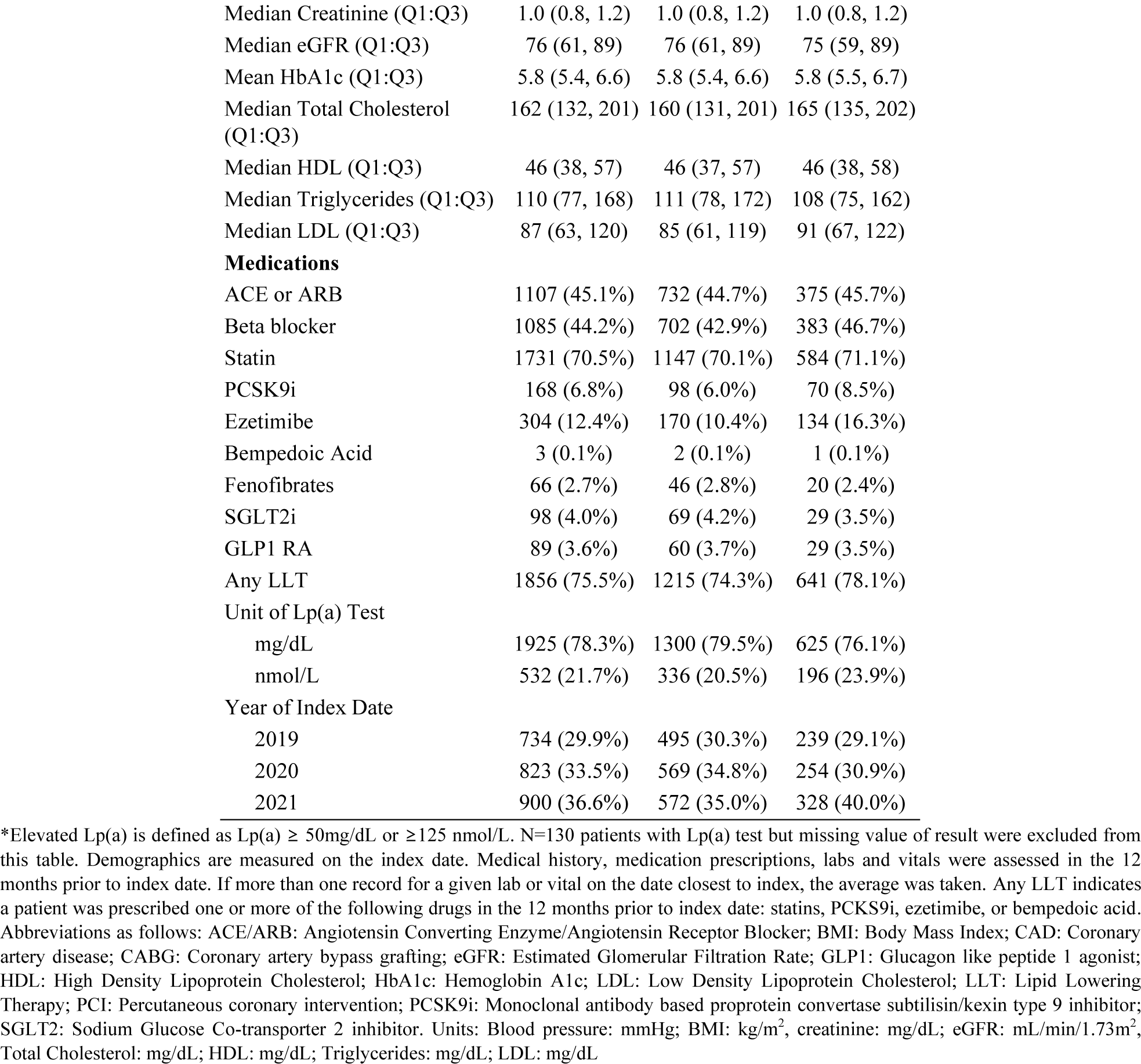
Baseline Characteristics based on Elevated Lp(a) status*.

**Supplementary table 2:** Initiation of Lipid Lowering Therapy by Lp(a) level.

| LLT | Lp(a)<50<br>mg/dL | 50-100 mg/dL | Lp(a)>100<br>mg/dL | p-value |
| --- | --- | --- | --- | --- |
| N in group | N=1300 | N=374 | N=251 |  |
| Any LLT | 985 (75.8%) | 286 (76.5%) | 208 (82.9%) |  |
| Initiated Statins | 117/368 (31.8%) | 42/120 (35.0%) | 16/54 (29.6%) | 0.734 |
| Initiated High Intensity Statins | 104/706 (14.7%) | 40/214 (18.7%) | 15/114 (13.2%) | 0.292 |
| Initiated PCSK9 | 68/1227 (5.5%) | 25/338 (7.4%) | 36/233 (15.5%) | <.001 |
| Initiated Ezetimibe | 84/1151 (7.3%) | 41/321 (12.8%) | 30/192 (15.6%) | <.001 |
| Initiated Bempedoic Acid | 3/1299 (0.2%) | 0/373 (0.0%) | 0/251 (0.0%) | 0.846 |
| Initiated Any LLT | 128/315 (40.6%) | 44/88 (50.0%) | 18/43 (41.9%) | 0.290 |

Initiation defined by no prescription in the year prior to index date but a prescription is present in the 6 months following an Lp(a) test done in units of mg/dL (N=1,925). Lp(a) level defined as <50mg/dL, 50-100mg/dL, and >100mg/dL Any LLT indicates a patient was initiated on one or more of the following prescriptions: statins, PCSK9i, ezetimibe, or bempedoic acid. For patients on high dose statins and PCSK9i, the proportion of patients on other LLTs prior to their initiation are listed. LLT: lipid lowering therapy; PCSK9i: Monoclonal antibody based proprotein convertase subtilisin/kexin type 9 inhibitor

**Supplementary table 3:** Multivariable regression analysis of likelihood of testing for Lp(a)

| **Factor** | **Relative Risk**<br>**(95% CI)** | **P-value** |
| --- | --- | --- |
| Age (yrs) | 0.75 (0.73-0.76) | <.001 |
| Female vs male | 0.96 (0.89-1.05) | 0.408 |
| Race (ref: white) |  | <.001 |
| Black | 0.76 (0.67-0.87) | <.001 |
| Other | 0.98 (0.80-1.19) | 0.817 |
| Hispanic vs non-Hispanic | 0.84 (0.65-1.09) | 0.195 |
| SBP (mmHg) | 0.98 (0.97-0.99) | 0.002 |
| DBP (mmHg) | 1.04 (1.02-1.06) | <.001 |
| BMI | 0.89 (0.86-0.91) | <.001 |
| Current smoker vs former/never | 0.45 (0.39-0.51) | <.001 |
| CAD | 1.78 (1.58-1.99) | <.001 |
| Ischemic stroke or TIA | 1.42 (1.29-1.57) | <.001 |
| PAD | 1.39 (1.27-1.53) | <.001 |
| Heart failure | 1.24 (1.13-1.37) | <.001 |
| Atrial fibrillation | 1.08 (0.97-1.20) | 0.184 |
| Hypertension | 0.69 (0.62-0.77) | <.001 |
| Hyperlipidemia | 1.58 (1.38-1.81) | <.001 |
| Diabetes | 0.68 (0.61-0.77) | <.001 |
| Hypertriglyceridemia | 1.07 (0.90-1.27) | 0.472 |
| Familial hypercholesterolemia | 1.44 (1.25-1.66) | <.001 |
| CKD | 1.02 (0.91-1.14) | 0.766 |
| Ace/ARB | 1.04 (0.95-1.14) | 0.360 |
| Beta blocker | 0.74 (0.68-0.81) | <.001 |
| Lipid lowering therapy (ref: none) |  | <.001 |
| Statin and non-statin LLT | 3.62 (3.14-4.19) | <.001 |
| Non-statin LLT only | 3.28 (2.73-3.94) | <.001 |
| High-intensity statin only | 1.65 (1.46-1.85) | <.001 |
| Moderate or low intensity statin only | 1.21 (1.07-1.38) | 0.003 |
| HbA1c (ref: < 6.0%) |  | <.001 |
| Missing | 0.56 (0.51-0.62) | <.001 |
| 6.5-7.9% | 0.81 (0.70-0.95) | 0.009 |
| 8-10% | 0.81 (0.66-1.00) | 0.046 |
| >10% | 0.85 (0.65-1.11) | 0.236 |
| LDL (ref: < 70 mg/dL) |  | <.001 |
| Missing | 0.47 (0.34-0.64) | <.001 |
| 70-99 mg/dL | 0.90 (0.82-1.00) | 0.055 |
| 100-129 mg/dL | 1.00 (0.89-1.13) | 0.955 |
| 130-159 mg/dL | 1.17 (1.01-1.36) | 0.031 |
| 160-189 mg/dL | 1.68 (1.40-2.02) | <.001 |
| 190+ mg/dL | 2.29 (1.84-2.83) | <.001 |
| HDL (ref: 41-59 mg/dL) |  | <.001 |
| Missing | 0.49 (0.27-0.90) | 0.022 |
| ≤40 mg/dL | 0.83 (0.75-0.91) | <.001 |
| ≥60 mg/dL | 1.13 (1.01-1.26) | 0.027 |
| Triglycerides (ref: < 150 mg/dL) |  | <.001 |

| Factor | Relative Risk<br>(95% CI) | P-value |
| --- | --- | --- |
| Missing | 0.36 (0.20-0.65) | <.001 |
| 150-199 mg/dL | 0.90 (0.80-1.02) | 0.093 |
| 200-249 mg/dL | 1.03 (0.91-1.16) | 0.640 |
| 250-500 mg/dL | 1.49 (1.04-2.13) | 0.029 |
| eGFR (ref: >= 90) |  | 0.004 |
| Missing | 1.30 (1.07-1.59) | 0.009 |
| <60 | 0.96 (0.83-1.11) | 0.539 |
| 60-89 | 1.11 (0.99-1.23) | 0.065 |

**Supplementary Table 4:** Diagnosis codes used for analyses.

| DIAGNOSIS | CODE TYPE | CODES |
| --- | --- | --- |
| ATRIAL FIBRILLATION | DX09 | 427.31 |
| ATRIAL FIBRILLATION | DX10 | I48.0, I48.1, I48.11, I48.19, I48.2, I48.20, I48.21, I48.91 |
| STABLE ANGINA | DX09 | 413.0, 413.9 |
| STABLE ANGINA | DX10 | I20, I20.1, I20.8, I20.9 |
| UNSTABLE ANGINA | DX09 | 411.1 |
| UNSTABLE ANGINA | DX10 | I20.0 |
| AORTIC VALVE STENOSIS | DX09 | 395.0, 395.2, 396.0, 396.2, 746.3, 747.22 |
| AORTIC VALVE STENOSIS | DX10 | I06.0, I06.2, I35.0, I35.2, Q23.0, Q24.4, Q25.3 |
| CORONARY<br>REVASCULARIZATION, CABG | DX09 | V45.81 |
| CORONARY<br>REVASCULARIZATION, CABG | DX10 | Z95.1 |
| CHRONIC KIDNEY DISEASE | DX09 | 285.21, 403, 403.0, 403.00, 403.01, 403.1, 403.10, 403.11, 403.9, 403.90, 403.91, 404, 404.0, 404.00, 404.01, 404.02, 404.03, 404.1, 404.10, 404.11, 404.12, 404.13, 404.9, 404.90, 404.91, 404.92, 404.93, 585, 585.1, 585.2, 585.3, 585.4, 585.5, 585.6, 585.9 |
| CHRONIC KIDNEY DISEASE | DX10 | D63.1, E08.22, E09.22, E10.22, E11.22, E13.22, I12, I12.0, I12.9, I13, I13.0, I13.1, I13.10, I13.11, I13.2, N18, N18.1, N18.2, N18.3, N18.30, N18.31, N18.32, N18.4, N18.5, N18.6, N18.9 |
| DIABETES MELLITUS | DX09 | 250, 250.0, 250.00, 250.01, 250.02, 250.03, 250.1, 250.10, 250.11, 250.12, 250.13, 250.2, 250.20, 250.21, 250.22, 250.23, 250.3, 250.30, 250.31, 250.32, 250.33, 250.4, 250.40, 250.41, 250.42, 250.43, 250.5, 250.50, 250.51, 250.52, 250.53, 250.6, 250.60, 250.61, 250.62, 250.63, 250.7, 250.70, 250.71, 250.72, 250.73, 250.8, 250.80, 250.81, 250.82, 250.83, 250.9, 250.90, 250.91, 250.92, 250.93 |
| DIABETES MELLITUS | DX10 | E10, E10.1, E10.10, E10.11, E10.2, E10.21, E10.22, E10.29, E10.3, E10.31, E10.311, E10.319, E10.32, E10.321, E10.3211, E10.3212, E10.3213, E10.3219, E10.329, E10.3291, E10.3292, E10.3293, E10.3299, E10.33, E10.331, E10.3311, E10.3312, E10.3313, E10.3319, E10.339, E10.3391, E10.3392, E10.3393, E10.3399, E10.34, E10.341, E10.3411, E10.3412, E10.3413, E10.3419, E10.349, E10.3491, E10.3492, E10.3493, E10.3499, E10.35, E10.351, E10.3511, E10.3512, E10.3513, E10.3519, E10.352, E10.3521, E10.3522, E10.3523, E10.3529, E10.353, E10.3531, E10.3532, E10.3533, E10.3539, E10.354, E10.3541, E10.3542, E10.3543, E10.3549, E10.355, E10.3551, E10.3552, E10.3553, E10.3559, E10.359, E10.3591, E10.3592, E10.3593, E10.3599, E10.36, E10.37, E10.37X1, E10.37X2, E10.37X3, E10.37X9, E10.39, E10.4, E10.40, E10.41, E10.42, E10.43, E10.44, E10.49, E10.5, E10.51, E10.52, E10.59, E10.6, E10.61, E10.610, E10.618, E10.62, E10.620, E10.621, E10.622, E10.628, E10.63, E10.630, E10.638, E10.64, E10.641, E10.649, E10.65, E10.69, E10.8, E10.9, E11, E11.0, E11.00, E11.01, E11.1, E11.10, E11.11, E11.2, E11.21, E11.22, E11.29, E11.3, E11.31, E11.311, E11.319, E11.32, E11.321, E11.3211, E11.3212, E11.3213, E11.3219, E11.329, E11.3291, E11.3292, E11.3293, E11.3299, E11.33, E11.331, E11.3311, E11.3312, E11.3313, E11.3319, E11.339, E11.3391, E11.3392, E11.3393, E11.3399, E11.34, E11.341, E11.3411, E11.3412, E11.3413, E11.3419, E11.349, E11.3491, E11.3492, E11.3493, E11.3499, E11.35, E11.351, E11.3511, E11.3512, E11.3513, E11.3519, E11.352, E11.3521, E11.3522, E11.3523, E11.3529, E11.353, E11.3531, E11.3532, E11.3533, E11.3539, E11.354, E11.3541, E11.3542, E11.3543, E11.3549, E11.355, E11.3551, E11.3552, E11.3553, E11.3559, E11.359, E11.3591, E11.3592, E11.3593, E11.3599, E11.36, E11.37, E11.37X1, E11.37X2, E11.37X3, E11.37X9, E11.39, E11.4, E11.40, E11.41, E11.42, E11.43, E11.44, E11.49, E11.5, E11.51, E11.52, E11.59, E11.6, E11.61, E11.610, E11.618, E11.62, E11.620, E11.621, E11.622, E11.628, E11.63, E11.630, E11.638, E11.64, E11.641, E11.649, E11.65, E11.69, E11.8, E11.9, E13, E13.0, E13.00, E13.01, E13.1, E13.10, E13.11, E13.2, E13.21, E13.22, E13.29, E13.3, E13.31, E13.311, E13.319, E13.32, E13.321, E13.3211, E13.3212, E13.3213, E13.3219, E13.329, E13.3291, E13.3292, E13.3293, E13.3299, E13.33, E13.331, E13.3311, E13.3312, E13.3313, E13.3319, E13.339, E13.3391, |
|  |  | E13.3392, E13.3393, E13.3399, E13.34, E13.341, E13.3411, E13.3412, E13.3413, E13.3419, E13.349, E13.3491, E13.3492, E13.3493, E13.3499, E13.35, E13.351, E13.3511, E13.3512, E13.3513, E13.3519, E13.352, E13.3521, E13.3522, E13.3523, E13.3529, E13.353, E13.3531, E13.3532, E13.3533, E13.3539, E13.354, E13.3541, E13.3542, E13.3543, E13.3549, E13.355, E13.3551, E13.3552, E13.3553, E13.3559, E13.359, E13.3591, E13.3592, E13.3593, E13.3599, E13.36, E13.37, E13.37X1, E13.37X2, E13.37X3, E13.37X9, E13.39, E13.4, E13.40, E13.41, E13.42, E13.43, E13.44, E13.49, E13.5, E13.51, E13.52, E13.59, E13.6, E13.61, E13.610, E13.618, E13.62, E13.620, E13.621, E13.622, E13.628, E13.63, E13.630, E13.638, E13.64, E13.641, E13.649, E13.65, E13.69, E13.8, E13.9 |
| FAMILIAL<br>HYPERCHOLESTEROLEMIA | DX09 | 272.0 |
| FAMILIAL<br>HYPERCHOLESTEROLEMIA | DX10 | E78.01 |
| HEART FAILURE | DX09 | 398.91, 402.01, 402.11, 402.91, 404.01, 404.03, 404.11, 404.13, 404.91, 404.93, 428, 428.0, 428.1, 428.2, 428.20, 428.21, 428.22, 428.23, 428.3, 428.30, 428.31, 428.32, 428.33, 428.4, 428.40, 428.41, 428.42, 428.43, 428.9 |
| HEART FAILURE | DX10 | I09.81, I11.0, I13.0, I13.2, I50, I50.1, I50.2, I50.20, I50.21, I50.22, I50.23, I50.3, I50.30, I50.31, I50.32, I50.33, I50.4, I50.40, I50.41, I50.42, I50.43, I50.8, I50.81, I50.810, I50.811, I50.812, I50.813, I50.814, I50.82, I50.83, I50.84, I50.89, I50.9 |
| HYPERLIPIDEMIA | DX09 | 272.0, 272.1, 272.2, 272.3, 272.4 |
| HYPERLIPIDEMIA | DX10 | E78.0, E78.00, E78.01, E78.1, E78.2, E78.3, E78.4, E78.41, E78.49, E78.5 |
| HYPERTENSION | DX09 | 401, 401.0, 401.1, 401.9, 402, 402.0, 402.00, 402.01, 402.1, 402.10, 402.11, 402.9, 402.90, 402.91, 403, 403.0, 403.00, 403.01, 403.1, 403.10, 403.11, 403.9, 403.90, 403.91, 404, 404.0, 404.00, 404.01, 404.02, 404.03, 404.1, 404.10, 404.11, 404.12, 404.13, 404.9, 404.90, 404.91, 404.92, 404.93, 405, 405.0, 405.01, 405.09, 405.1, 405.11, 405.19, 405.9, 405.91, 405.99 |
| HYPERTENSION | DX10 | I10, I11, I11.0, I11.9, I12, I12.0, I12.9, I13, I13.0, I13.1, I13.10, I13.11, I13.2, I15, I15.0, I15.1, I15.2, I15.8, I15.9 |
| HYPERTRIGLYCERIDEMIA | DX09 | 272.1 |
| HYPERTRIGLYCERIDEMIA | DX10 | E78.1 |
| ISCHEMIC HEART DISEASE | DX09 | 410.00, 410.01, 410.02, 410.10, 410.11, 410.12, 410.20, 410.21, 410.22, 410.30, 410.31, 410.32, 410.40, 410.41, 410.42, 410.50, 410.51, 410.52, 410.60, 410.61, 410.62, 410.70, 410.71, 410.72, 410.80, 410.81, 410.82, 410.90, 410.91, 410.92, 411.0, 411.1, 411.81, 411.89, 412, 413.0, 413.9, 414.00, 414.01, 414.02, 414.03, 414.04, 414.05, 414.06, 414.07, 414.12, 414.2, 414.3, 414.4, 414.8, 414.9 |
| ISCHEMIC HEART DISEASE | DX10 | I20.0, I20.8, I20.9, I21.01, I21.02, I21.09, I21.11, I21.19, I21.21, I21.29, I21.3, I21.4, I21.A1, I21.A9, I22.0, I22.1, I22.2, I22.8, I22.9, I23.0, I23.1, I23.2, I23.3, I23.4, I23.5, I23.6, I23.7, I23.8, I24.0, I24.1, I24.8, I24.9, I25.1, I25.10, I25.11, I25.110, I25.111, I25.118, I25.119, I25.2, I25.5, I25.6, I25.70, I25.700, I25.701, I25.708, I25.709, I25.71, I25.710, I25.711, I25.718, I25.719, I25.72, I25.720, I25.721, I25.728, I25.729, I25.73, I25.730, I25.731, I25.738, I25.739, I25.750, I25.751, I25.758, I25.759, I25.760, I25.761, I25.768, I25.769, I25.79, I25.790, I25.791, I25.798, I25.799, I25.8, I25.81, I25.810, I25.811, I25.812, I25.82, I25.83, I25.84, I25.89, I25.9 |
| MYOCARDIAL INFARCTION, EVENT | DX09 | 410.00, 410.01, 410.10, 410.11, 410.20, 410.21, 410.30, 410.31, 410.40, 410.41, 410.50, 410.51, 410.60, 410.61, 410.70, 410.71, 410.80, 410.81, 410.90, 410.91 |
| MYOCARDIAL INFARCTION, EVENT | DX10 | I21, I21.0, I21.01, I21.02, I21.09, I21.1, I21.11, I21.19, I21.2, I21.21, I21.29, I21.3, I21.4 |
| MYOCARDIAL INFARCTION, HISTORY | DX09 | 410, 410.0, 410.00, 410.01, 410.02, 410.1, 410.10, 410.11, 410.12, 410.2, 410.20, 410.21, 410.22, 410.3, 410.30, 410.31, 410.32, 410.4, 410.40, 410.41, 410.42, 410.5, 410.50, 410.51, 410.52, 410.6, 410.60, 410.61, 410.62, 410.7, 410.70, 410.71, 410.72, 410.8, 410.80, 410.81, 410.82, 410.9, 410.90, 410.91, 410.92, 412, 429.79 |
| MYOCARDIAL INFARCTION, HISTORY | DX10 | I21, I21.0, I21.01, I21.02, I21.09, I21.1, I21.11, I21.19, I21.2, I21.21, I21.29, I21.3, I21.4, I21.9, I21.A, I21.A1, I21.A9, I22, I22.0, I22.1, I22.2, I22.8, I22.9, I25.2 |
| PERIPHERAL ARTERIAL DISEASE | DX09 | 437.0, 440.0, 440.1, 440.2, 440.20, 440.21, 440.22, 440.23, 440.24, 440.29, 440.3, 440.30, 440.31, 440.32, 440.4, 443.2, 443.9, 557.1 |
| PERIPHERAL ARTERIAL DISEASE | DX10 | I65.2, I65.21, I65.22, I65.23, I65.29, I67.2, I70.0, I70.1, I70.2, I70.20, I70.201, I70.202, I70.203, I70.208, I70.209, I70.21, I70.211, I70.212, I70.213, I70.218, I70.219, I70.22, I70.221, I70.222, I70.223, I70.228, I70.229, I70.23, I70.231, I70.232, I70.233, I70.234, I70.235, I70.238, I70.239, I70.24, I70.241, I70.242, I70.243, I70.244, I70.245, I70.248, I70.249, I70.25, I70.26, I70.261, I70.262, I70.263, I70.268, I70.269, I70.29, I70.291, I70.292, I70.293, I70.298, I70.299, I70.3, I70.30, |
|  |  | I70.301, I70.302, I70.303, I70.308, I70.309, I70.31,<br>I70.311, I70.312, I70.313, I70.318, I70.319, I70.32,<br>I70.321, I70.322, I70.323, I70.328, I70.329, I70.33,<br>I70.331, I70.332, I70.333, I70.334, I70.335, I70.338,<br>I70.339, I70.34, I70.341, I70.342, I70.343, I70.344,<br>I70.345, I70.348, I70.349, I70.35, I70.36, I70.361,<br>I70.362, I70.363, I70.368, I70.369, I70.39, I70.391,<br>I70.392, I70.393, I70.398, I70.399, I70.4, I70.40, I70.401,<br>I70.402, I70.403, I70.408, I70.409, I70.41, I70.411,<br>I70.412, I70.413, I70.418, I70.419, I70.42, I70.421,<br>I70.422, I70.423, I70.428, I70.429, I70.43, I70.431,<br>I70.432, I70.433, I70.434, I70.435, I70.438, I70.439,<br>I70.44, I70.441, I70.442, I70.443, I70.444, I70.445,<br>I70.448, I70.449, I70.45, I70.46, I70.461, I70.462,<br>I70.463, I70.468, I70.469, I70.49, I70.491, I70.492,<br>I70.493, I70.498, I70.499, I70.5, I70.50, I70.501, I70.502,<br>I70.503, I70.508, I70.509, I70.51, I70.511, I70.512,<br>I70.513, I70.518, I70.519, I70.52, I70.521, I70.522,<br>I70.523, I70.528, I70.529, I70.53, I70.531, I70.532,<br>I70.533, I70.534, I70.535, I70.538, I70.539, I70.54,<br>I70.541, I70.542, I70.543, I70.544, I70.545, I70.548,<br>I70.549, I70.55, I70.56, I70.561, I70.562, I70.563,<br>I70.568, I70.569, I70.59, I70.591, I70.592, I70.593,<br>I70.598, I70.599, I70.6, I70.60, I70.601, I70.602, I70.603,<br>I70.608, I70.609, I70.61, I70.611, I70.612, I70.613,<br>I70.618, I70.619, I70.62, I70.621, I70.622, I70.623,<br>I70.628, I70.629, I70.63, I70.631, I70.632, I70.633,<br>I70.634, I70.635, I70.638, I70.639, I70.64, I70.641,<br>I70.642, I70.643, I70.644, I70.645, I70.648, I70.649,<br>I70.65, I70.66, I70.661, I70.662, I70.663, I70.668,<br>I70.669, I70.69, I70.691, I70.692, I70.693, I70.698,<br>I70.699, I70.7, I70.70, I70.701, I70.702, I70.703, I70.708,<br>I70.709, I70.71, I70.711, I70.712, I70.713, I70.718,<br>I70.719, I70.72, I70.721, I70.722, I70.723, I70.728,<br>I70.729, I70.73, I70.731, I70.732, I70.733, I70.734,<br>I70.735, I70.738, I70.739, I70.74, I70.741, I70.742,<br>I70.743, I70.744, I70.745, I70.748, I70.749, I70.75,<br>I70.76, I70.761, I70.762, I70.763, I70.768, I70.769,<br>I70.79, I70.791, I70.792, I70.793, I70.798, I70.799,<br>I70.92, I79.0, Z95.8, Z95.820, Z98.62 |
| CORONARY<br>REVASCULARIZATION, PCI | DX09 | V45.82 |
| CORONARY<br>REVASCULARIZATION, PCI | DX10 | Z95.5, Z98.61 |
| ISCHEMIC STROKE | DX09 | 433.01, 433.11, 433.21, 433.31, 433.81, 433.91, 434.01,<br>434.11, 434.91 |
| ISCHEMIC STROKE | DX10 | I63, I63.0, I63.00, I63.01, I63.011, I63.012, I63.013, I63.019, I63.02, I63.03, I63.031, I63.032, I63.033, I63.039, I63.09, I63.1, I63.10, I63.11, I63.111, I63.112, I63.113, I63.119, I63.12, I63.13, I63.131, I63.132, I63.133, I63.139, I63.19, I63.2, I63.20, I63.21, I63.211, I63.212, I63.213, I63.219, I63.22, I63.23, I63.231, I63.232, I63.233, I63.239, I63.29, I63.3, I63.30, I63.31, I63.311, I63.312, I63.313, I63.319, I63.32, I63.321, I63.322, I63.323, I63.329, I63.33, I63.331, I63.332, I63.333, I63.339, I63.34, I63.341, I63.342, I63.343, I63.349, I63.39, I63.4, I63.40, I63.41, I63.411, I63.412, I63.413, I63.419, I63.42, I63.421, I63.422, I63.423, I63.429, I63.43, I63.431, I63.432, I63.433, I63.439, I63.44, I63.441, I63.442, I63.443, I63.449, I63.49, I63.5, I63.50, I63.51, I63.511, I63.512, I63.513, I63.519, I63.52, I63.521, I63.522, I63.523, I63.529, I63.53, I63.531, I63.532, I63.533, I63.539, I63.54, I63.541, I63.542, I63.543, I63.549, I63.59, I63.8, I63.81, I63.89, I63.9 |
| TRANSIENT ISCHEMIC ATTACK | DX09 | 435, 435.0, 435.1, 435.2, 435.3, 435.8, 435.9 |
| TRANSIENT ISCHEMIC ATTACK | DX10 | G45, G45.0, G45.1, G45.2, G45.3, G45.4, G45.8, G45.9 |

**Supplementary Table 5:**
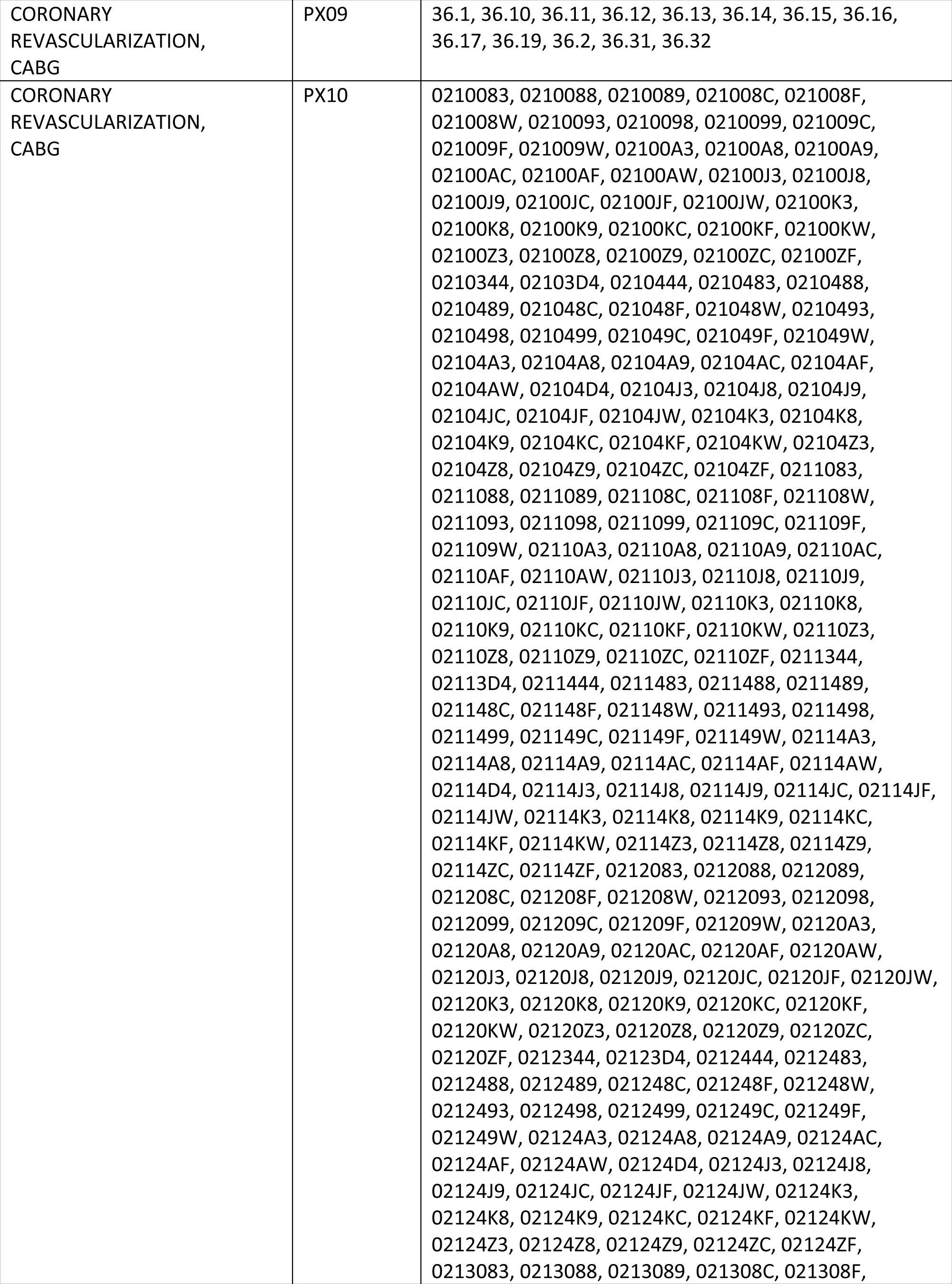
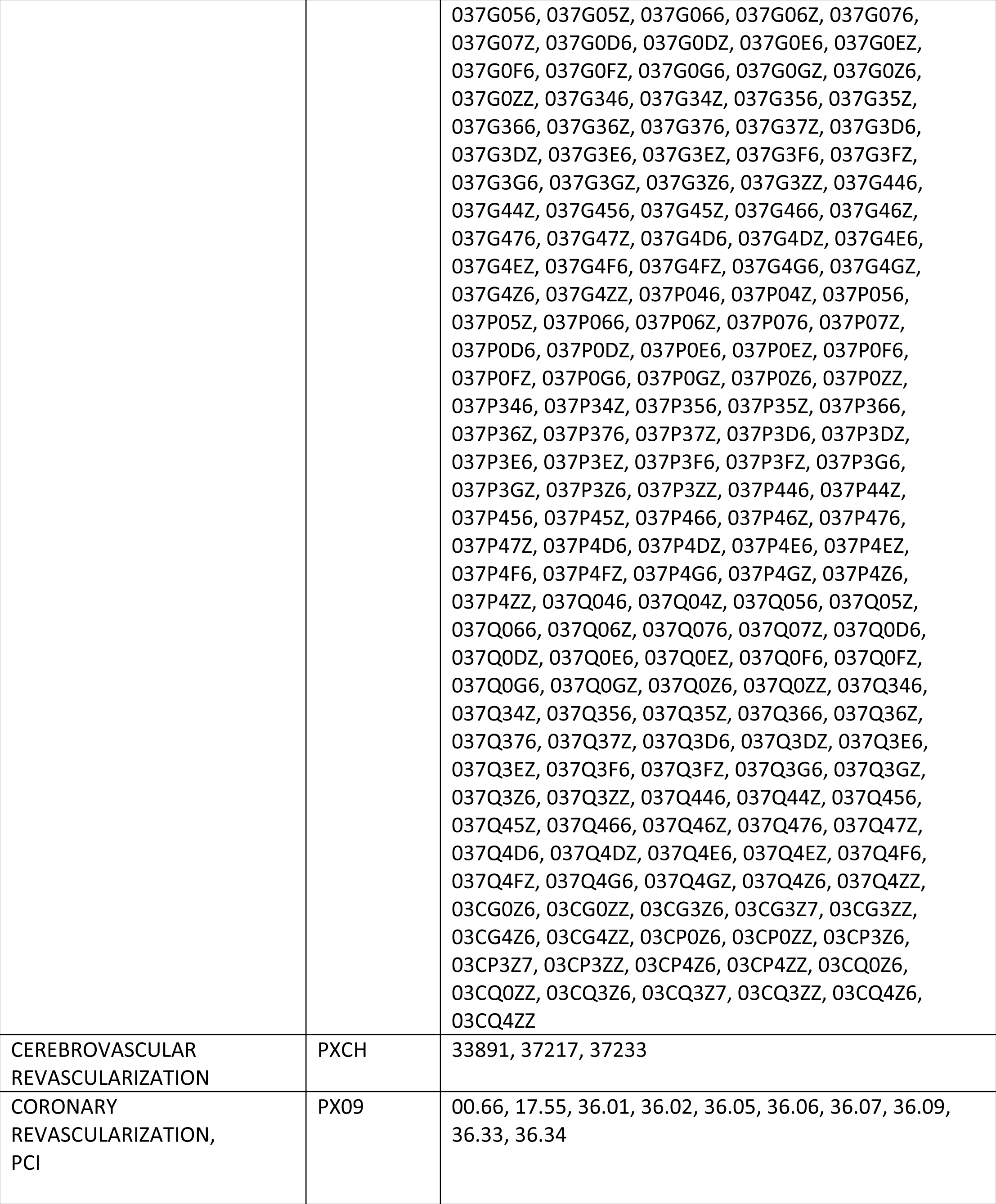
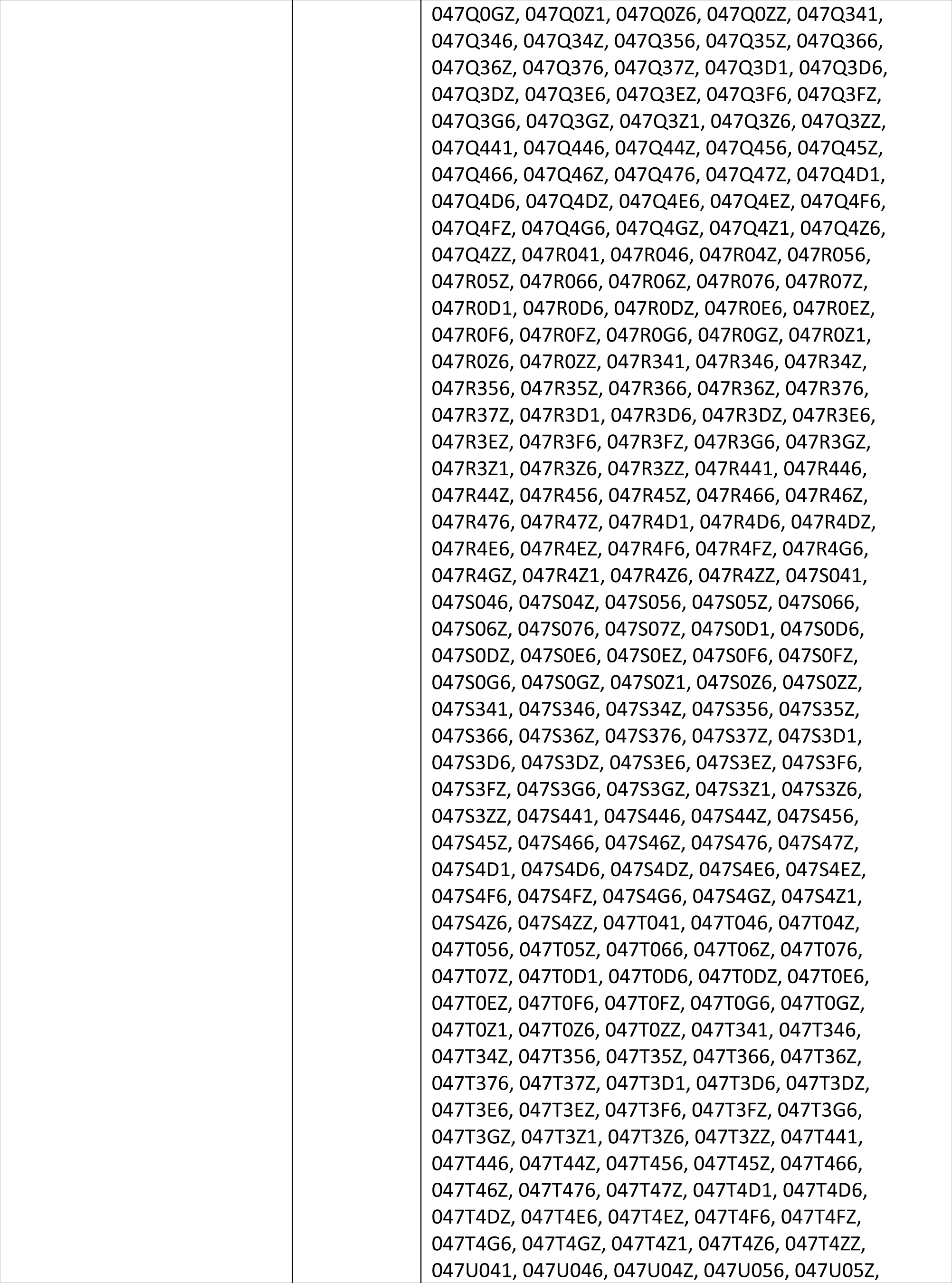
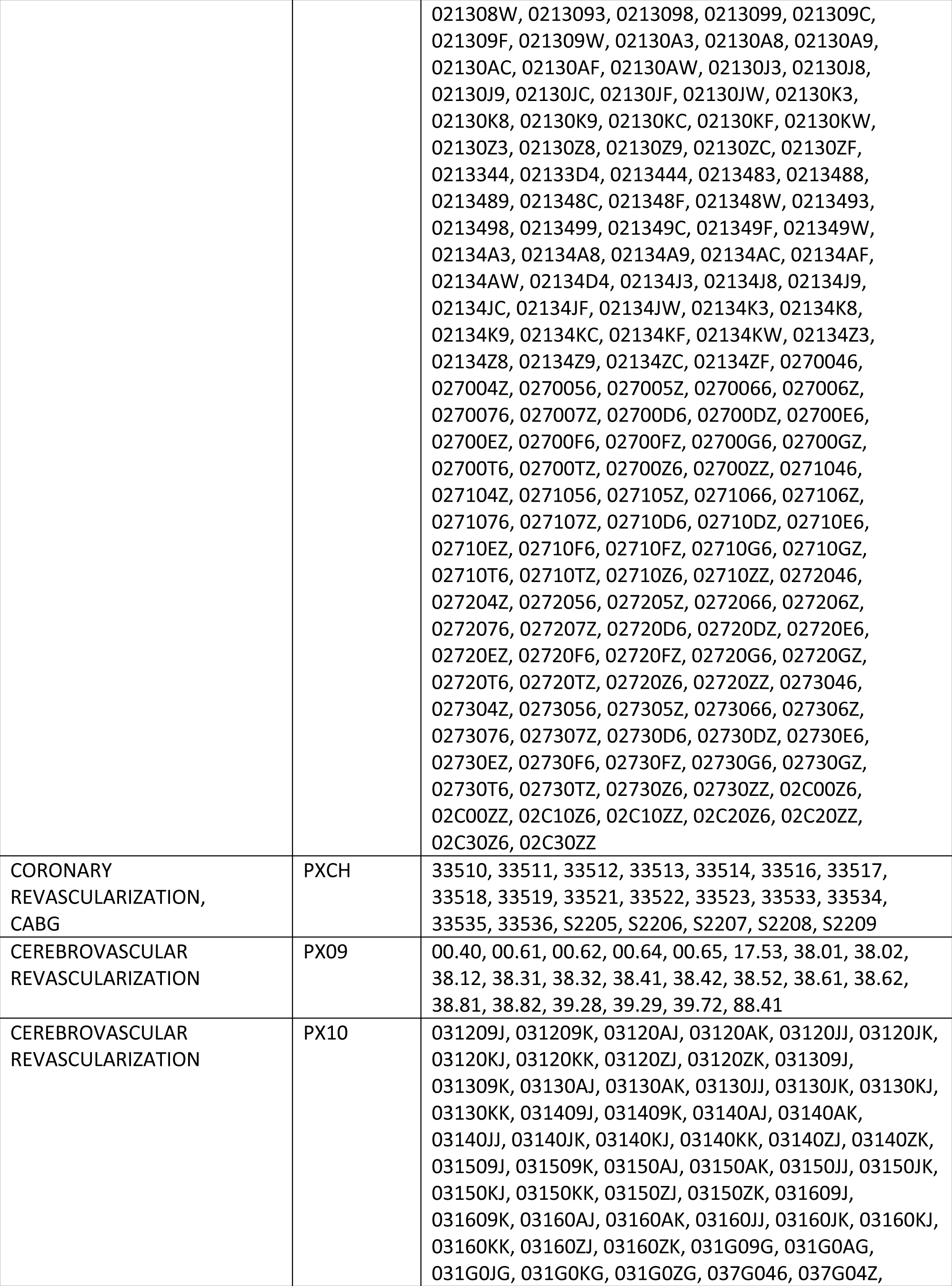
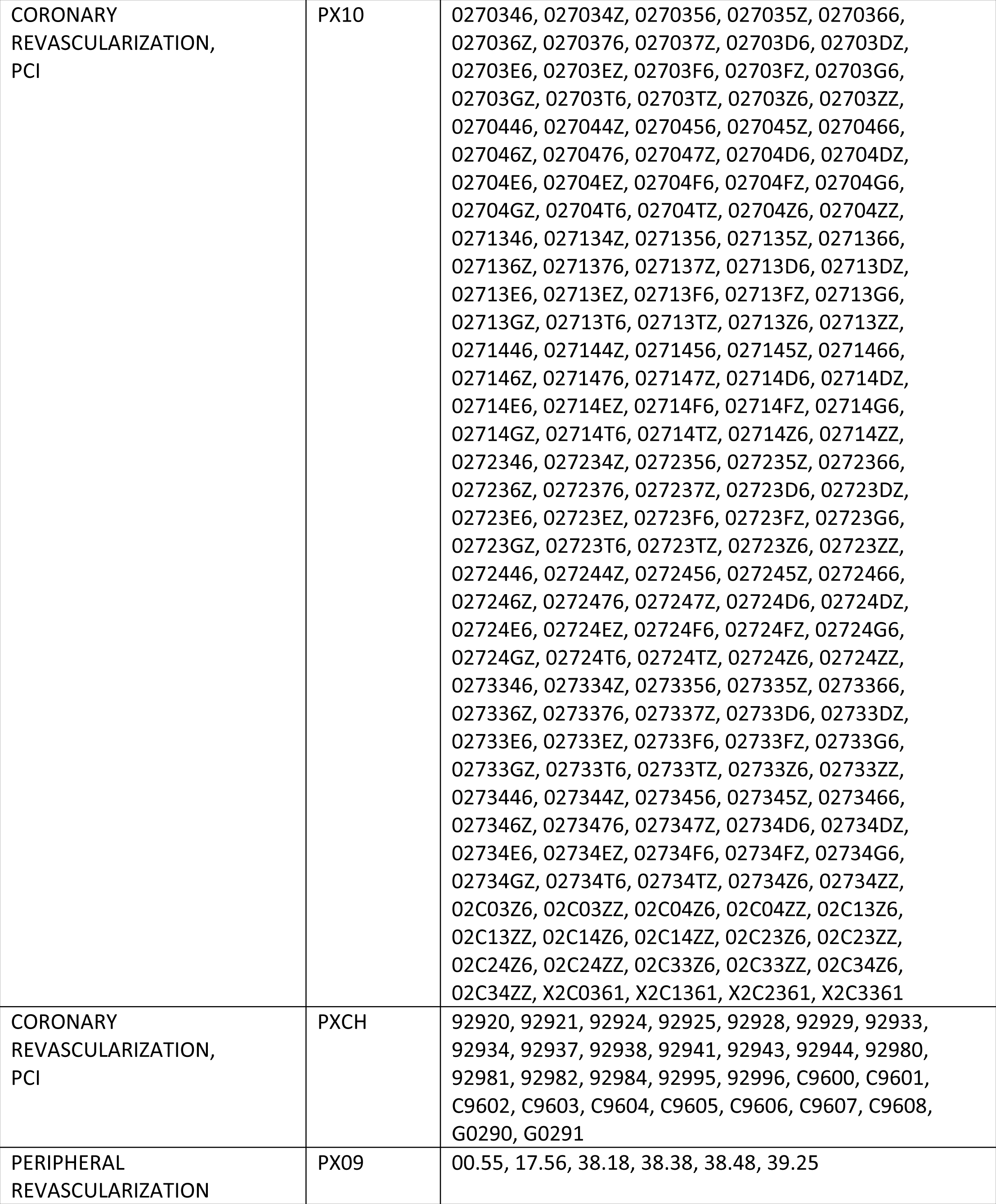
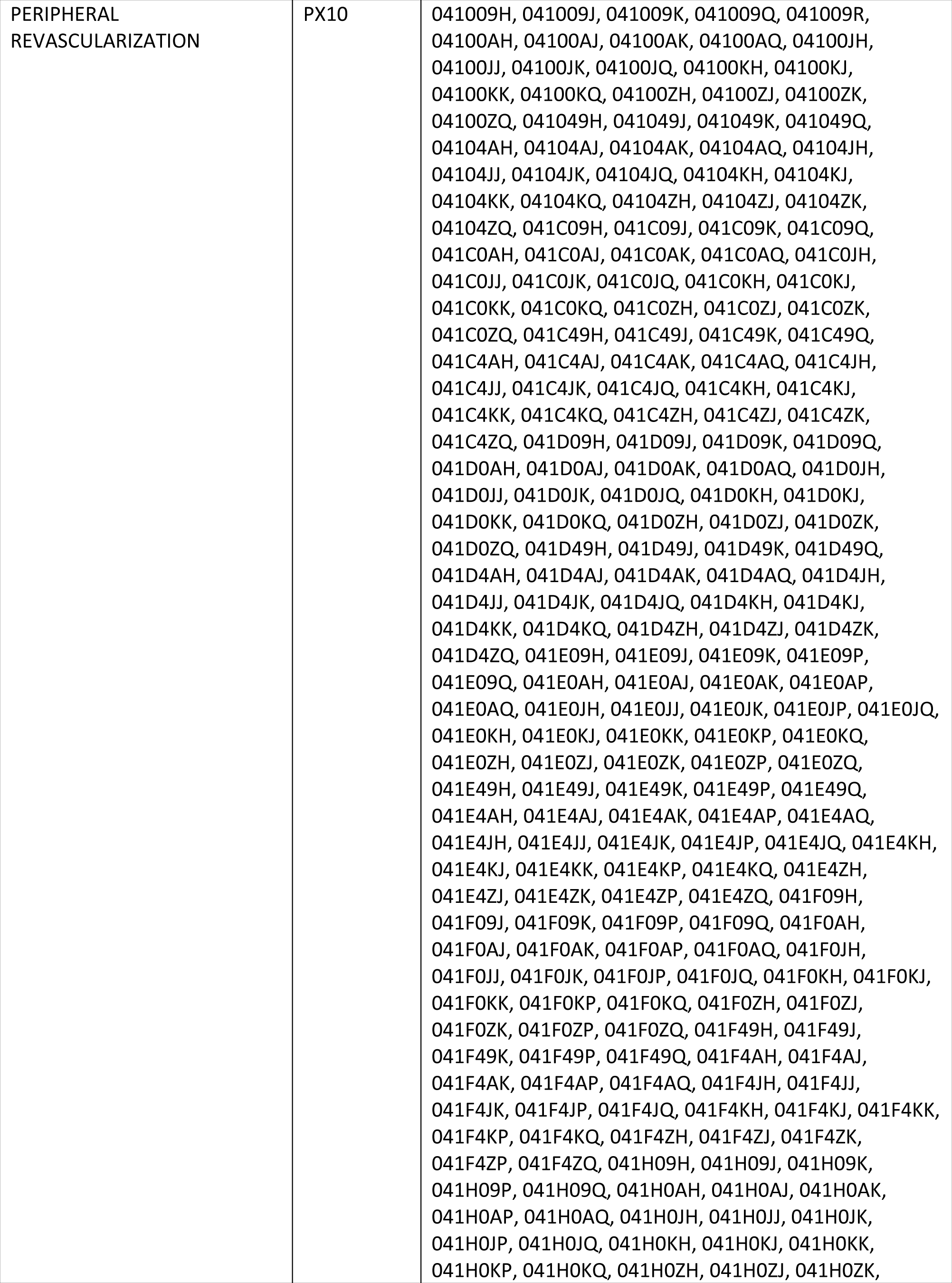
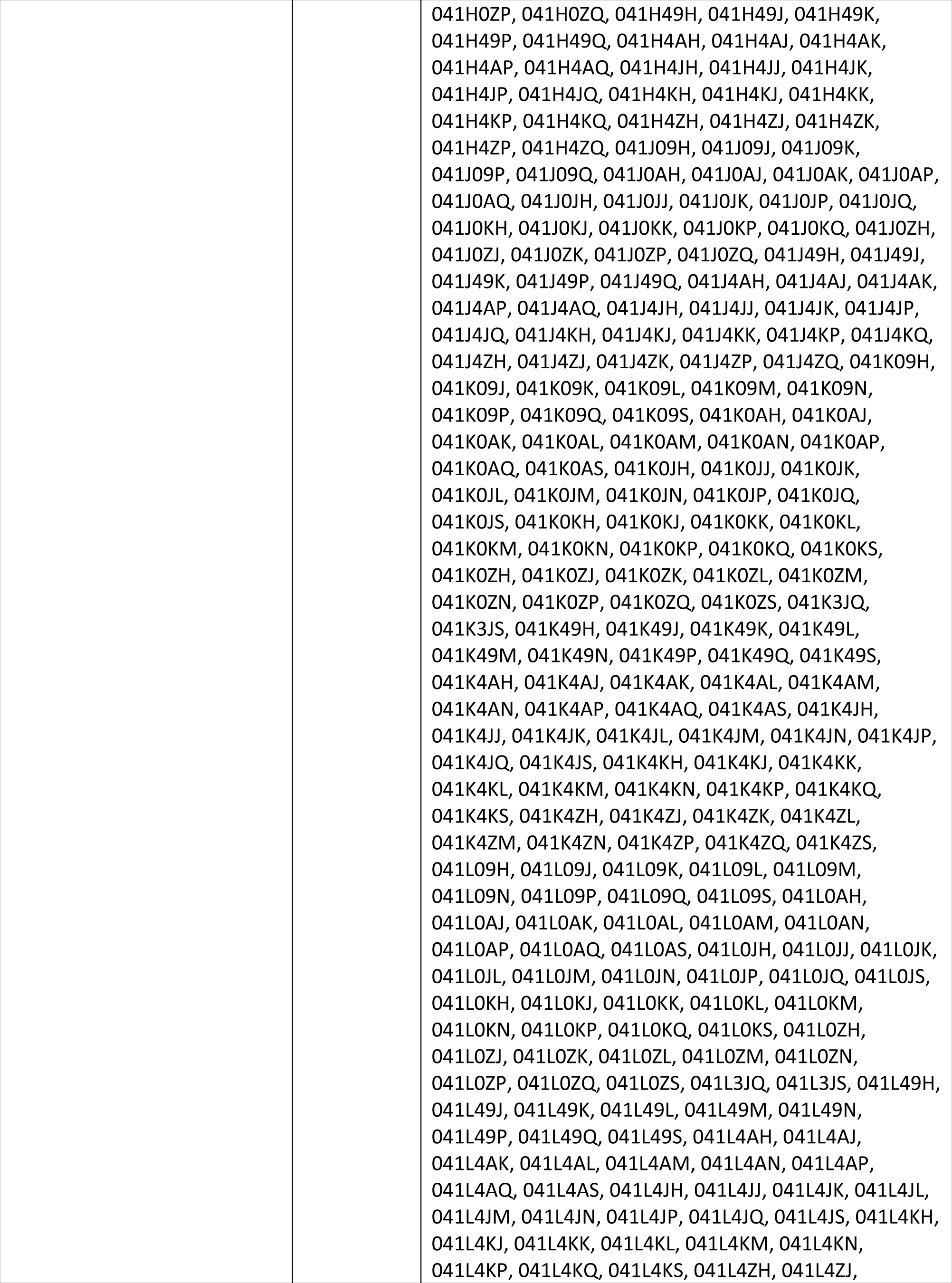
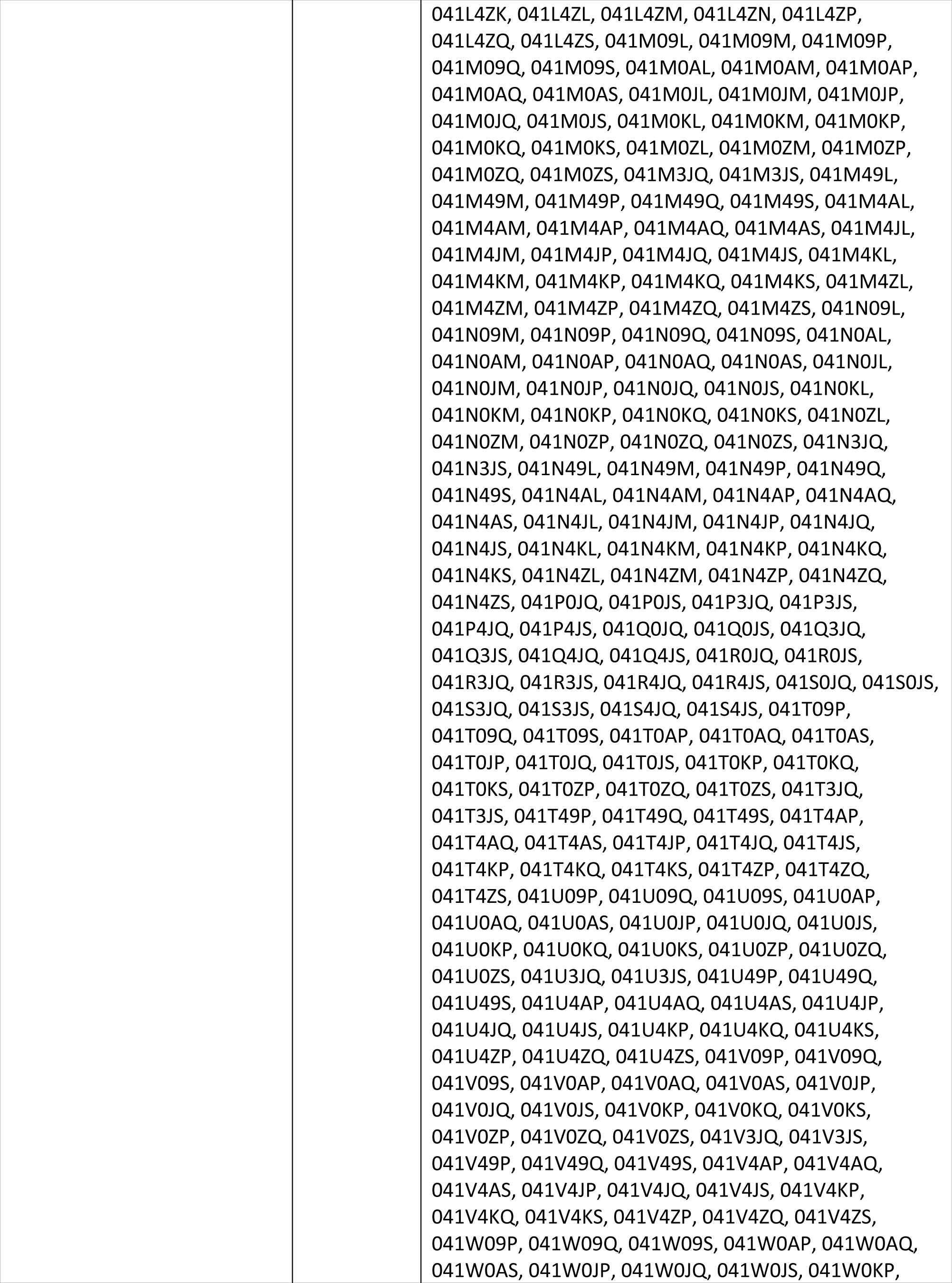
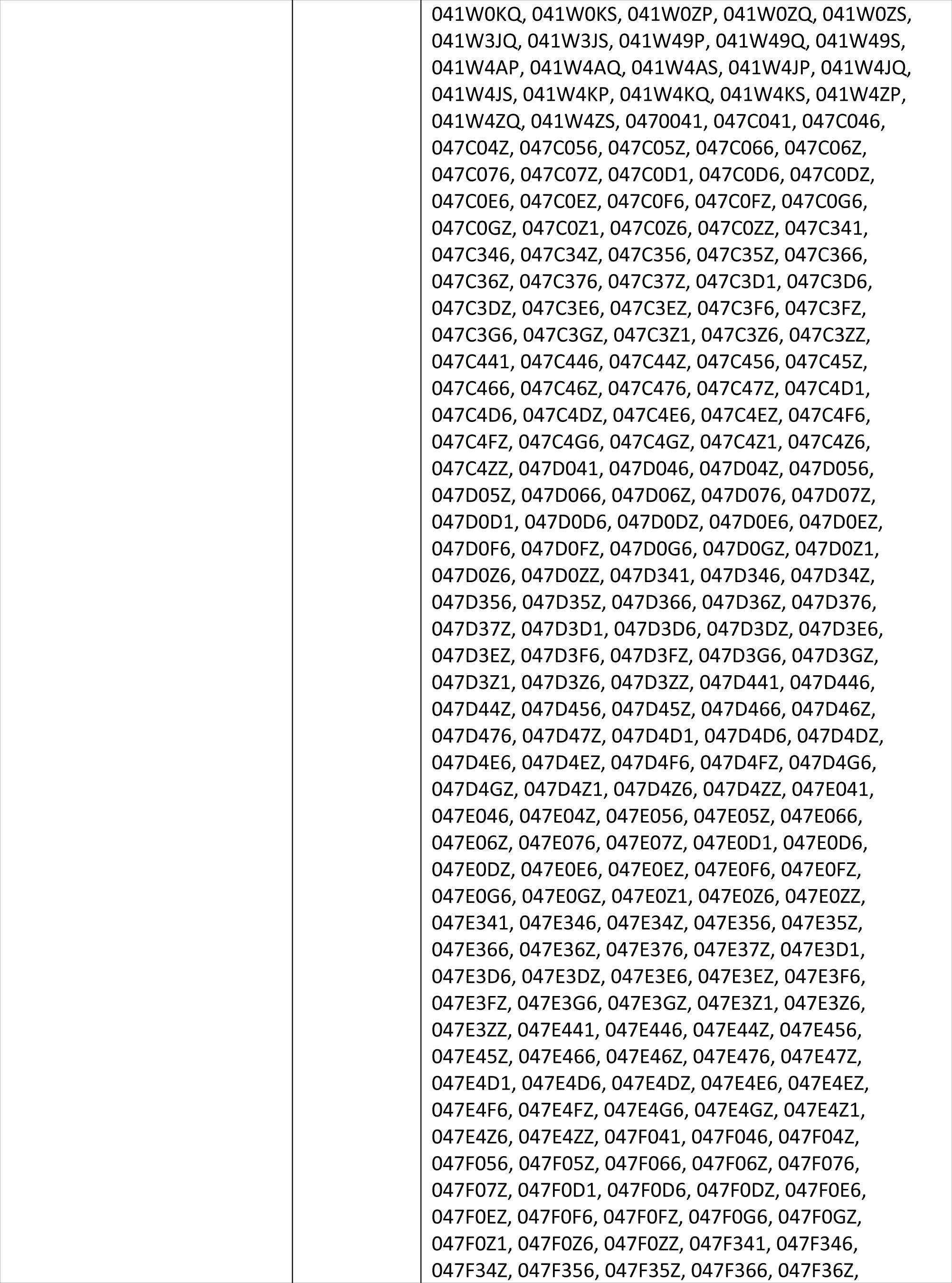
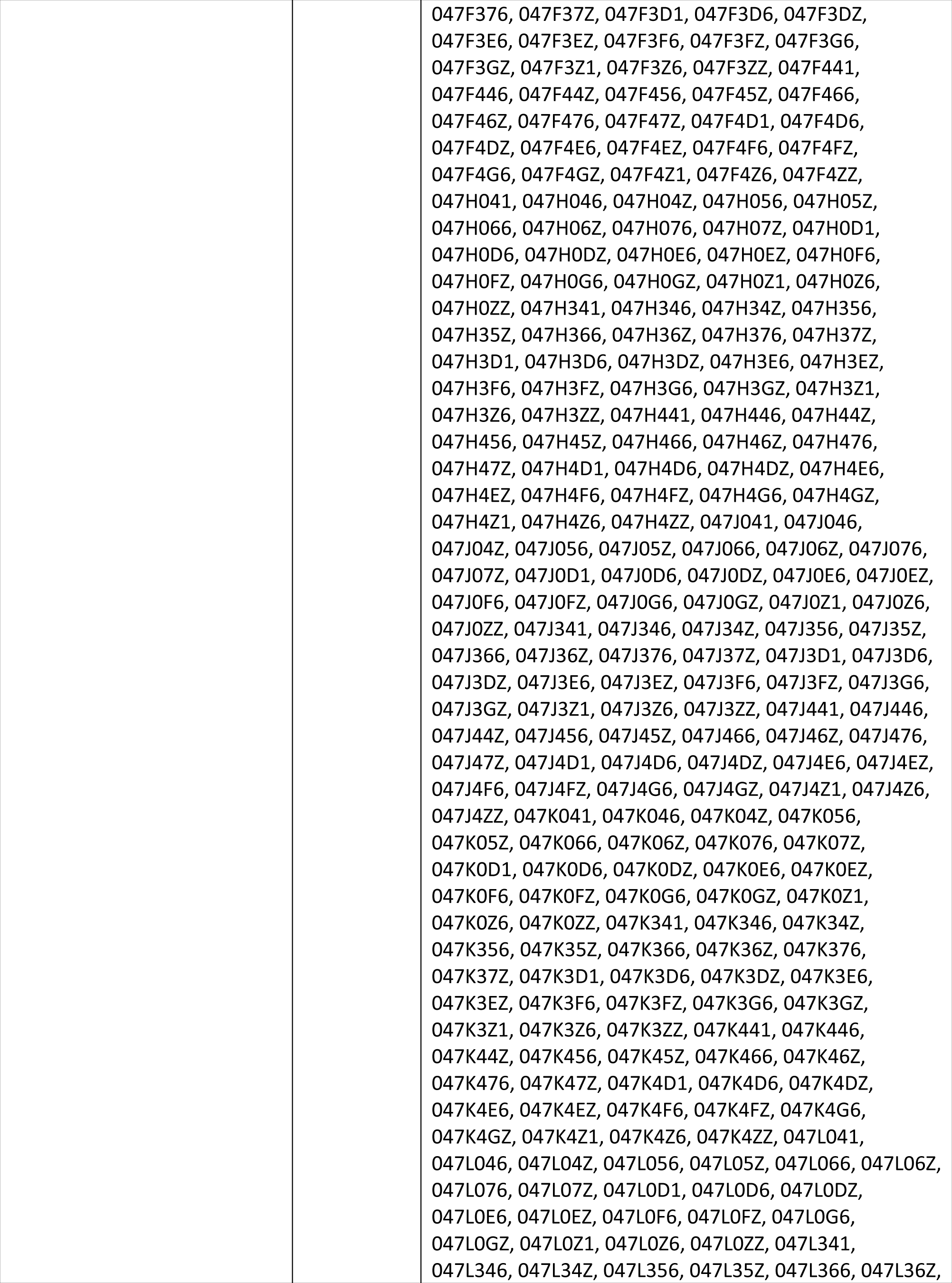
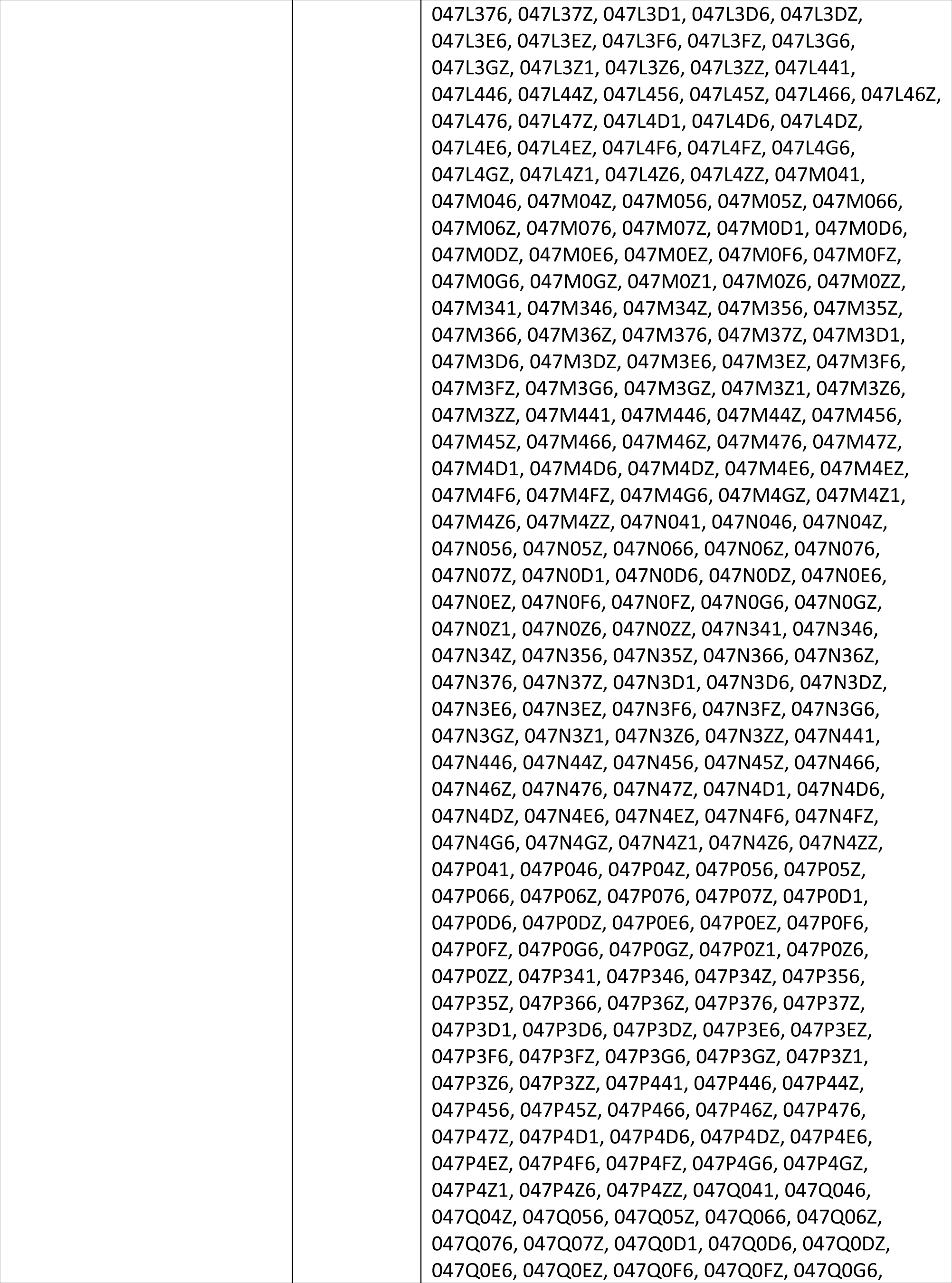
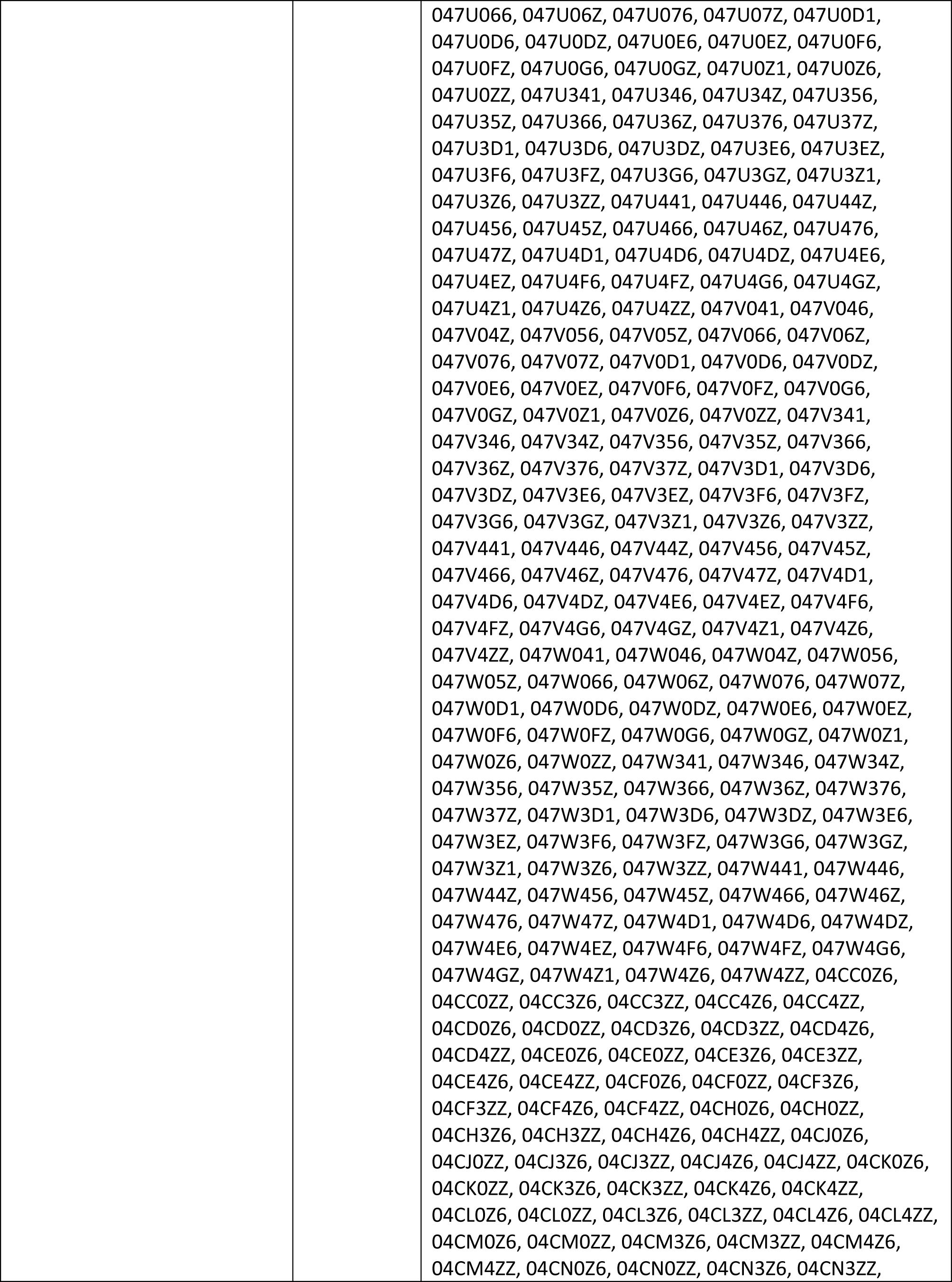
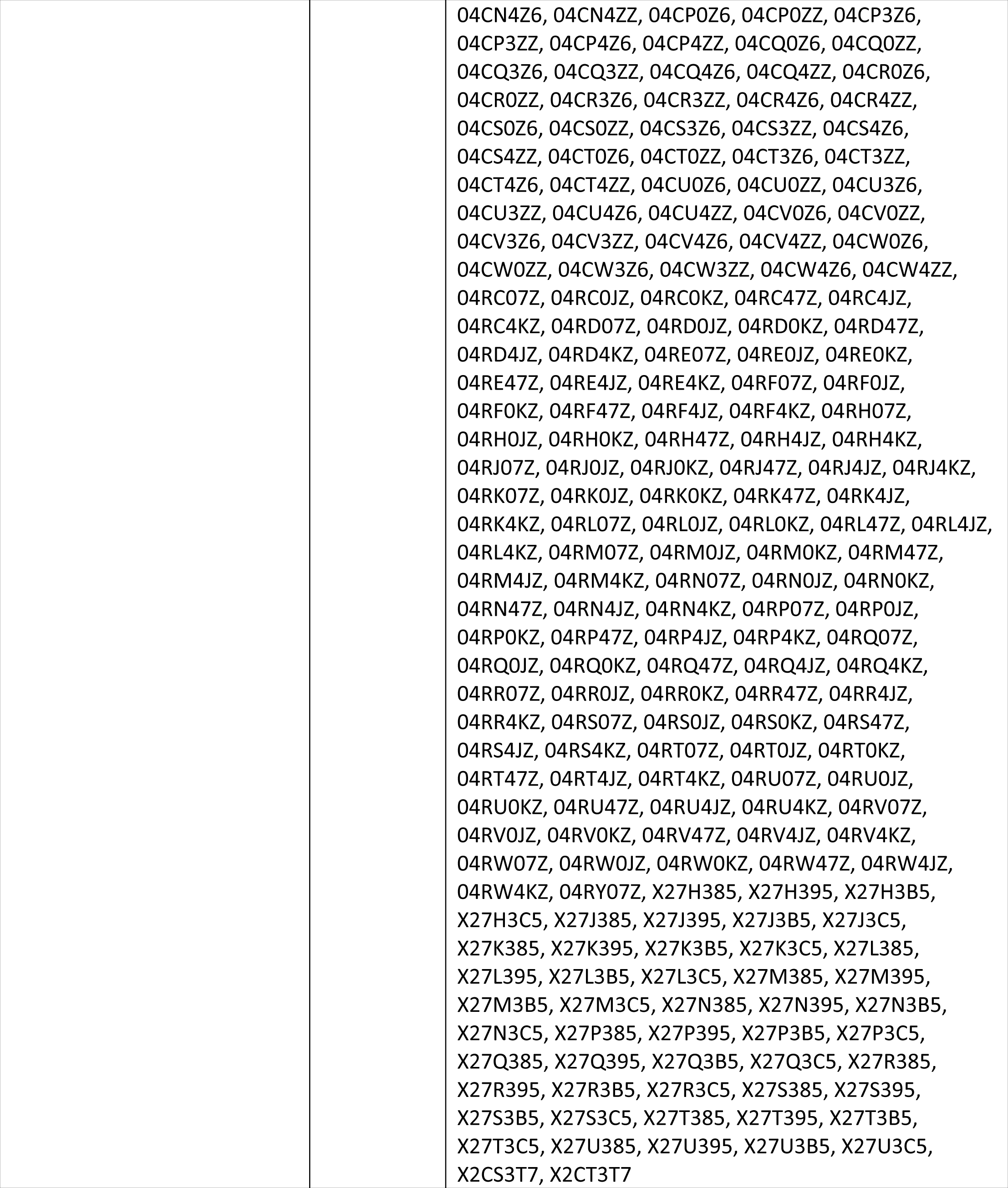
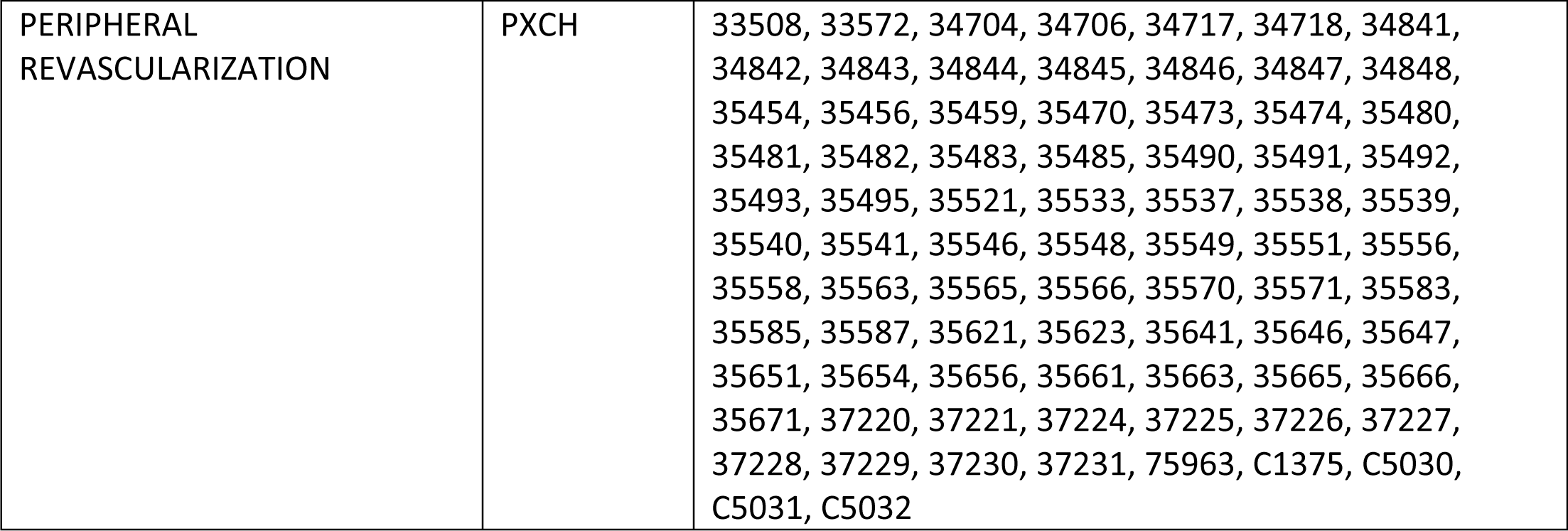
Procedure codes used for analysis.

## Notes

### Competing Interest Statement

The authors have declared no competing interest.

### Clinical Trial

N/A, this was not a clinical trial

### Funding Statement

Novartis Pharmaceuticals Corporation provided funding with a grant to support this study, including the principle investigators, corresponding author, and statistical support effort. The analytic plan, data analysis, data interpretation, or decision to publish as the study was investigator initiated with collaboration with the sponsor.

### Author Declarations

The study was approved by the Duke University institutional review board under a waiver of HIPAA authorization and informed consent

